# Ranking the Effectiveness of Non-Pharmaceutical Interventions to Counter COVID-19 in UK Universities with Vaccinated Population

**DOI:** 10.1101/2021.11.07.21266028

**Authors:** Zirui Niu, Giordano Scarciotti

**Author notes:** Contributing authors.

## Abstract

Several universities around the world have resumed in-person teaching after successful vaccination campaigns have covered 70/80% of the population. In this study, we combine a new compartmental model with an optimal control formulation to discover, among different non-pharmaceutical interventions, the best prevention strategy to maximize on-campus activities while keeping spread under control. Composed of two interconnected Susceptible-Exposed-Infected-Quarantined-Recovered (SEIQR) structures, the model enables staff-to-staff infections, student-to-staff cross infections, student-to-student infections, and environment-to-individual infections. Then, we model input variables representing the implementation of different non-pharmaceutical interventions and formulate and solve optimal control problems for four desired scenarios: minimum number of cases, minimum intervention, minimum non-quarantine intervention, and minimum quarantine intervention. Our results reveal the particular significance of mask wearing and social distancing in universities with vaccinated population (with proportions according to UK data). The study also reveals that quarantining infected students has a higher importance than quarantining staff. In contrast, other measures such as environmental disinfection seems to be less important.

---

The epidemic ascribed to the virus called Severe Acute Respiratory Syndrome Coronavirus 2 (SARS-CoV-2) has greatly impacted society and the economy around the world. In December 2019, China reported cases of pneumonia of unknown cause in Wuhan. The World Health Organisation (WHO) declared the coronavirus disease 2019 (COVID-19) a pandemic in March 11, 2020 [1]. By 3 November 2021, WHO has reported over 246 million cases with more than 5 million deaths around the world [1].

The virus generally causes respiratory symptoms such as cough, sneezing, shortness of breath, along with other symptoms including fever, headache [2] and olfactory or gustatory dysfunctions [3]. Since direct contact and aerosol transmissions are two important ways of infection [4], symptomatic individuals are high spreaders. Meanwhile, the virus can also survive on surfaces and then invade the human body through eyes, nose or mouth via touching [5, 6]. Some carriers are asymptomatic, but they can still infect other susceptible individuals [7, 8].

Many countries have conducted successful vaccination campaigns. However, vaccination uptake in most of these countries have platooned at around 70/80% of their total population [9]. Moreover, SARS-CoV-2 has shown a relatively high ability to adapt [10]. Several variants have been reported [11] with four considered to be of main concern: Alpha (B.1.1.7), Beta (B.1.351), Gamma (P.1) and Delta (B.1.617.2). In particular, the Delta variant has shown to be more infectious and more severe, leading to second or third waves in the UK, India and South Africa [12], in addition to having partial resistance to vaccines [13, 14]. The fast emergence of viral mutations has also raised wide considerations on whether current vaccines will be effective on new lineages appearing in the future [15–17]. Moreover, recent studies also show a decay of the protection that vaccines offer as time from vaccination increases [18, 19], which prompted several counties to start a booster campaign. Therefore, in such a complex situation the implementation of non-pharmaceutical interventions such as mask wearing and social distancing have remained fundamental measures in containing the disease [20].

This work studies the current situation in British universities which have re-opened and maintain a combination of non-pharmaceutical measures in place. The objective of universities is to maximise on-campus activities while maintaining the spread of the disease under control. Universities are “small-environments” which have special features for which general purpose models may be inadequate. For instance, a university is composed of two fundamentally different populations, the students and the staff, which have different degrees of interaction, vaccination rates and serious symptoms. General purpose models focus mostly on modelling the spread of the disease, but here we are interested in maximising on-campus activity subject to limited spread. Our main contribution is two-fold: on one hand we provide a modelling framework to maximise safe on-campus activity. On the other hand, a ranking of non-pharmaceutical interventions and their fundamental importance in achieving the objective naturally emerges from our analysis. Moreover this emergent behaviour is shown to be relative robust to modelling parameters.

The first block of our framework is a compartmental model. Compartmental modelling is a popular choice for research on COVID-19. Cooper et al. [21] used an SIR model with changing total population to estimate the growth of the epidemic in different nations. To explicitly model details such as incubation period, hospitalization, and quarantine, Leontitsis et al. [22] proposed an SEAHIR model. Giordano et al. [23] created a far more complete SIDARTHE model, reflecting potential effects of non-pharmaceutical interventions implemented by the government. Various stochastic versions with higher complexity have also been designed to estimate the development of the pandemic under feasible countermeasures [24–26]. However, most of these models consider the influence of prevention and control measures by tweaking the model’s parameter values. Consequently, the importance of different interventions cannot be systematically evaluated and compared. In this paper, we propose a new deterministic compartmental model for the spreading of COVID-19 in universities and investigate the importance of non-pharmaceutical countermeasures using optimal control techniques.

Consisting of a double SEIQR structure, our model distinguishes the epidemic evolution stages of students from those of staff. All individuals can potentially undergo five statuses: susceptible S (including the vaccinated), exposed E (asymptomatic), infected I (symptomatic), quarantined Q (hospitalized or isolated), and recovered R. Since all members are studying or working within the same confined space, the coronavirus can also transmit via the environment. An extra compartment C is used to represent the environmental virus concentration. The overall structure of the model is depicted in Fig. 1. This compartmental model automatically assumes that the total population is homogeneously mixed, which is reasonable because everyone belonging to the same department is following similar daily routines in the common confined spaces. The individuals in the compartments also have similar probability of infections due to the virus surviving in the environment. The model does not involve the isolation of asymptomatic subjects because it is less likely to occur in the context of a university population. We still model the vaccinated individuals as susceptible but with lower infection rates (*i*.*e*. vaccine breakthrough infections). The possibility of re-infection among recovered people is omitted. Death of vaccinated individuals is also negligible.

**Fig. 1.**
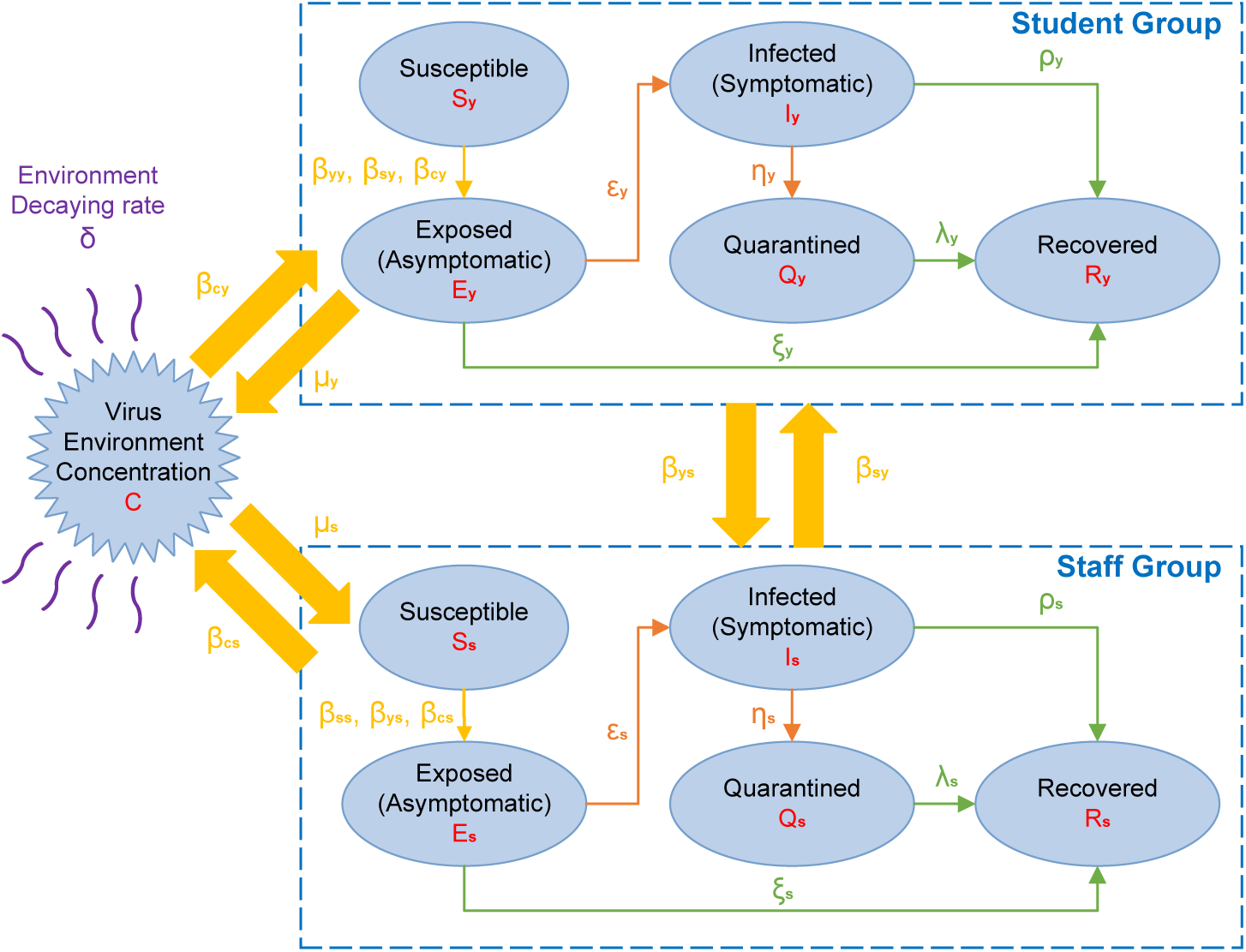
Model structure. Flow chart of five epidemic stages among students and staff in a university department: S, susceptible (including the vaccinated); E, exposed (asymptomatic); I, infected (symptomatic); Q, quarantined (hospitalized or mandatorily isolated); R, recovered. The subscript “y” stands for students while the subscript “s” denotes staff.

To assess the efficacy of the various intervention strategies under study we first evaluate a baseline scenario in which no countermeasures are implemented. The parameters for this scenario are taken from the literature. Some of the values of the parameters are obtained according to the proportion of vaccinated individuals and vaccine effectiveness reported in the UK [27–29]. A detailed description of this procedure is reported in the Methods.

We consider five possible non-pharmaceutical interventions that can be implemented by the university after reopening: mask wearing, social distancing, environmental disinfection, quarantine on infected students and quarantine of infected staff. To study the effectiveness of these measures in the model, we define five input control variables associated with each of these measures. We then use optimal control to study four desired scenarios: minimum number of cases, minimum intervention, minimum non-quarantine intervention and minimum quarantine intervention. In all scenarios, it is assumed that the University starts to control the epidemic 14 days after the asymptomatic carriers firstly appear. We also model the desire of keeping the infection under control as constraints in the optimisation. In particular, we impose constraints on the number of infected cases and on the number of days required to extinguish the epidemic. In the minimum-case scenario, the epidemic is controlled with no efforts spared, leading to the strongest minimization of COVID-19 cases. For minimum intervention, we study the possibility of minimizing the total effort of all control measures. In the minimum non-quarantine intervention, we minimise the use of mask wearing, social distancing and environmental disinfection. In the last scenario, minimum quarantine intervention, we minimize the use of quarantines. By comparing the obtained optimal trajectories in different scenarios, we identify an emergent behaviour that shows a ranking on the importance of the different non-pharmaceutical interventions.

## 1 Results

### 1.1 Baseline: no interventions

We first generate a baseline scenario in which no intervention is performed. We simulate the evolution of the epidemic in one department of the university. The results are shown in Fig. 2. As case study we consider a department where the total number of students is 1200 and the number of members of staff is 150 (i.e. similar to the EEE department of Imperial College London). Initially, we assume 5 students and 2 staff members start as asymptomatic cases. The effective reproduction number *R*_0_ on day 0 is 1.40, which indicates a potential outbreak of COVID-19 in this small environment. After two weeks, 1.8% of students and 3.1% of staff have caught the disease. Without countermeasures, increasingly more individuals will get infected during the coming 120 days. Simultaneously, *R*_*t*_ keeps decreasing and becomes smaller than 1 after 76 days. At day 134, 56% of students and 63% of staff have been infected and *R*_*t*_ = 0.718. In the end (without considering further infections from outside the department), the epidemic last around 250 days, rendering 59% of students and 66% of staff infected. This prediction result reveals the necessity of imposing non-pharmaceutical measures at the current UK vaccination levels (see Methods for the exact percentages).

**Fig. 2.**
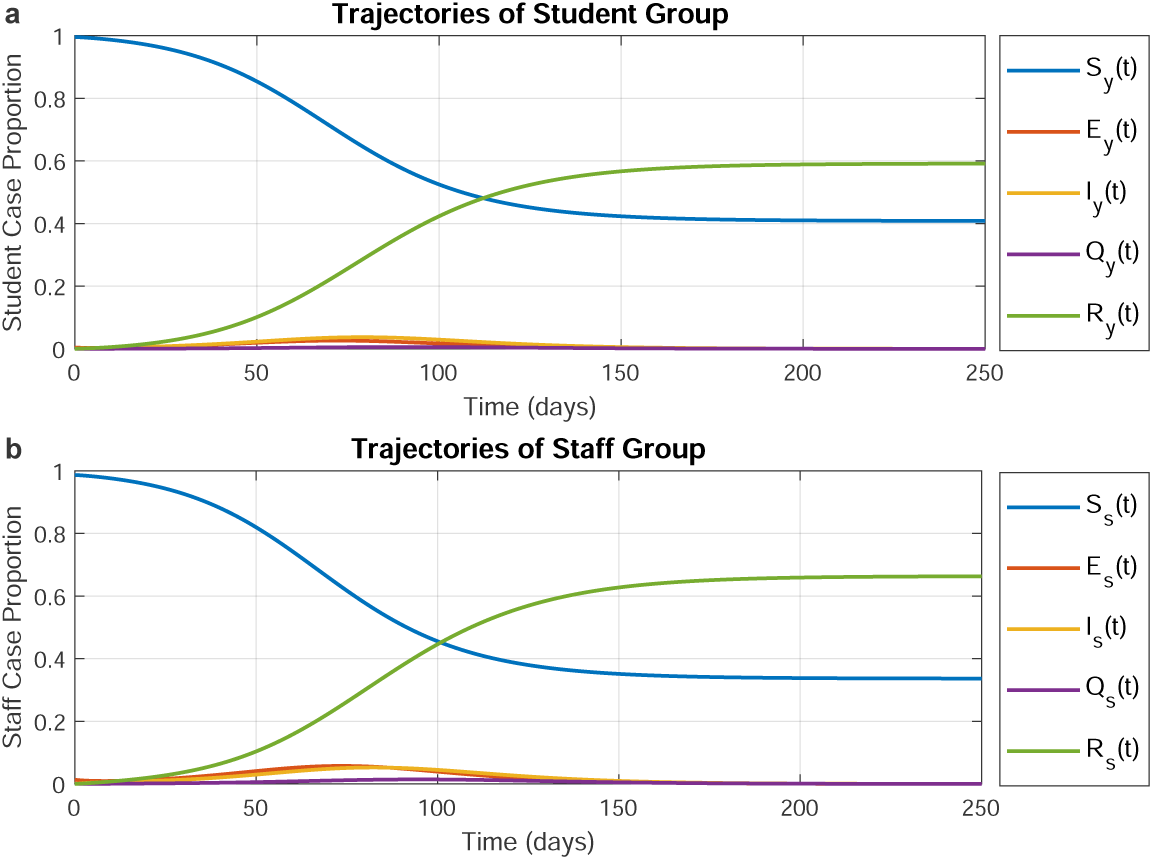
Prediction in the baseline scenario of no interventions. **a** Evolution of COVID-19 among students. **b** Evolution of COVID-19 among staff. Magnitudes are in proportion to the total number of students or staff.

### 1.2 Optimal interventions for four objectives

We now study the effect of non-pharmaceutical interventions. Which interventions to implement, when and how strongly are all decisions made by the optimisation method to optimise the objective. We have selected four different optimisation objectives. In all scenarios, interventions are introduced 14 days after the initial exposure of 5 students and 2 staff members. In all scenarios, we impose the constraints that the epidemic must end within 120 days and that at least 94% of students and staff are not infected. Thus, the overall timeline is 134 days.

#### Minimum number of cases

In this scenario the optimisation objective is formulated to minimise infections, even though this may require that all non-pharmaceutical interventions are implemented at full strength. The optimal trajectories are depicted in Fig. 3. The epidemic is completely ended at around the 60^th^ day when the individuals are only in two states: susceptible and recovered. More than 97% of students and 96% of staff do not get infected in this scenario. From the figure we see that this result is achieved by implementing all interventions unreservedly, from mask wearing to mandatory quarantine. This reduces *R*_*t*_ to around 0.217. While initially there is a strong need for all countermeasures, after 30 days the optimal strategy relies mostly on distancing and masks. All cases have been quarantined by the 45^th^ day. After approximately 60 days, the department reaches a steady state and the infection is stopped. Consequently, at this point the optimal strategy eases the interventions because the epidemic has been successfully contained.

**Fig. 3.**
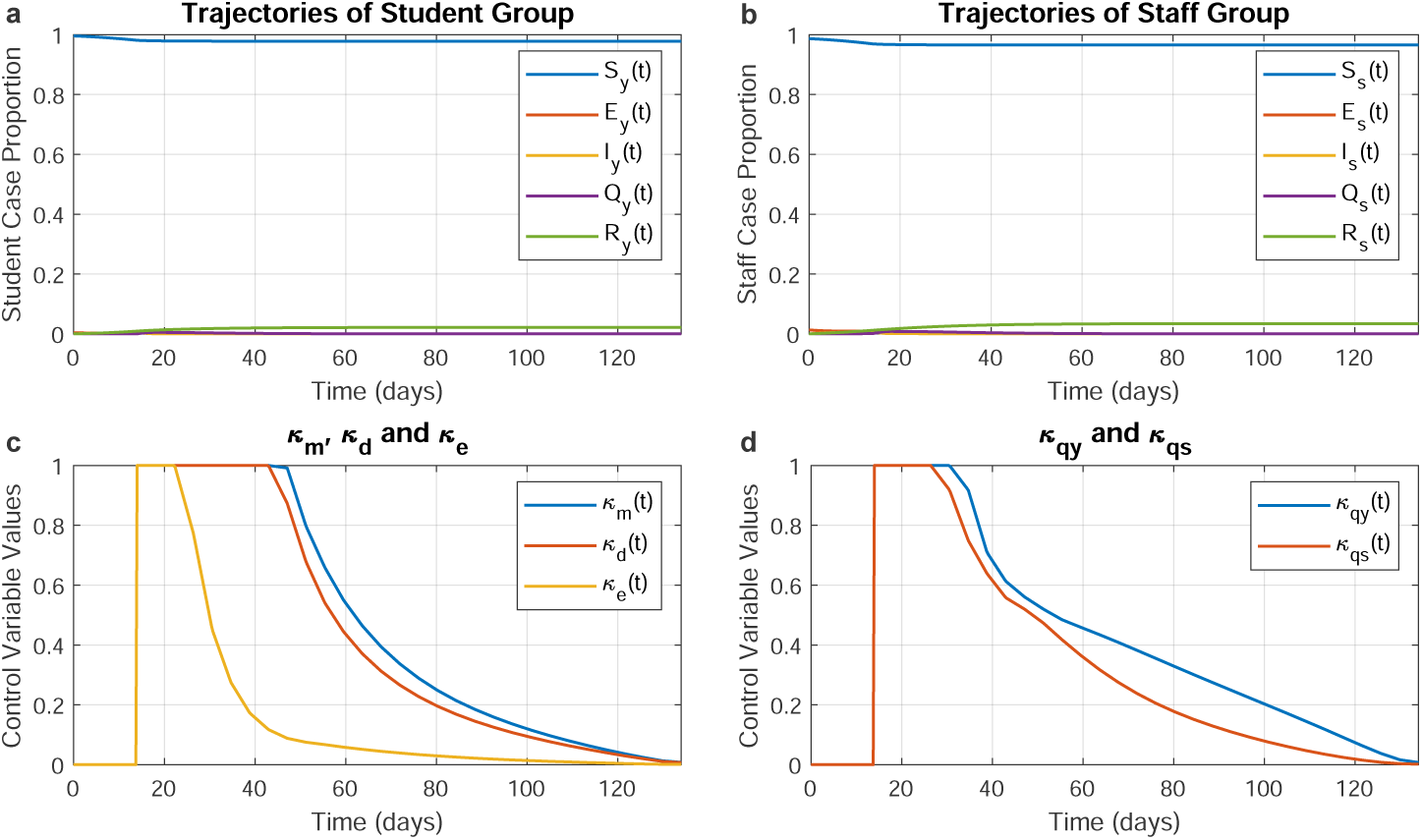
Optimal trajectories for the minimum-case scenario. Optimal trajectories when the department spares no effort to contain the epidemic. **a**,**b**, The epidemic evolution among students and staff, respectively. **c**, The optimal strategies for mask wearing (*κ*_*m*_), social distancing (*κ*_*d*_), and environmental disinfection (*κ*_*e*_). **d**, The optimal strategies for mandatory quarantine on infected students (*κ*_*qy*_) and infected staff (*κ*_*qs*_).

#### Minimum intervention

In this case the objective function is the norm of all control variables. As a consequence, the aim here is to minimise the use of all interventions (including quarantine) while still satisfying the constraint that at least 94% of the population is not infected. The resulting optimal trajectories are shown in Fig. 4. The epidemic is ended with 4.3% of students and 6% of staff having been infected. With respect to before we can see that there is a decrease in the strength of the interventions. We can also note that there is an emerging ranking between the interventions. Fig. 4c shows that the environmental disinfection is far less important than mask wearing. For what concerns mandatory quarantine shown in Fig. 4d, we notice again that isolation of infected students plays a more significant role in controlling the spread of COVID-19 than the isolation of staff. All five control interventions are strongest at the beginning of the epidemic, and then their magnitude is attenuated gradually over time.

**Fig. 4.**
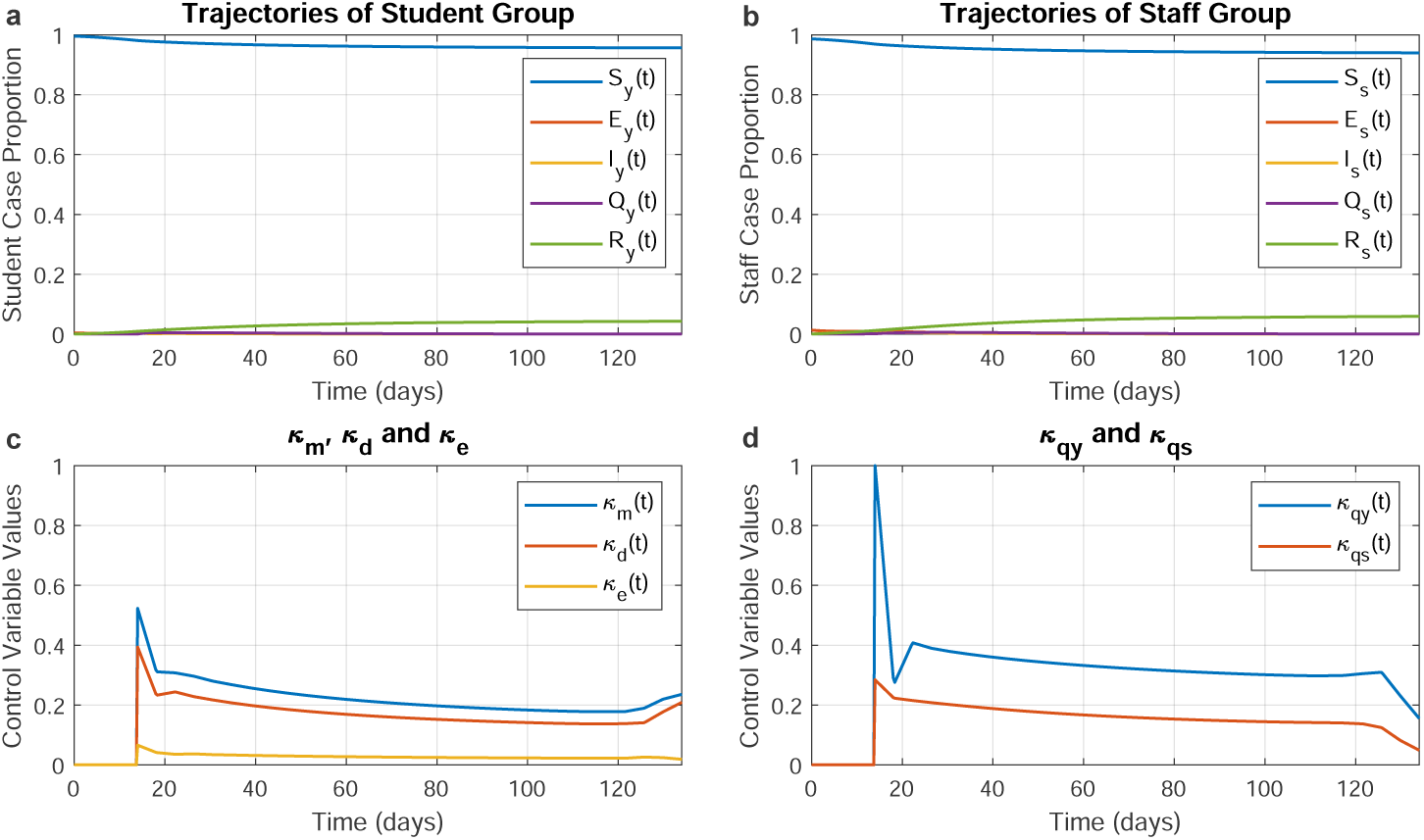
Optimal trajectories for the minimum intervention scenario. Optimal trajectories when the department would like to minimise the enforcement of control measures. **a**,**b**, The epidemic evolution among students and staff, respectively. **c**, The optimal strategies for mask wearing (*κ*_*m*_), social distancing (*κ*_*d*_), and environmental disinfection (*κ*_*e*_). **d**, The optimal strategies for mandatory quarantine on infected students (*κ*_*qy*_) and infected staff (*κ*_*qs*_).

#### Minimum use of non-quarantine interventions

In this case we want to minimize the use of masks, social distancing and environmental disinfection. As a result we expect an increase of the use of quarantines. The resulting optimal trajectories are shown in Fig. 5. As expected the figures show little use of non-quarantine interventions, and a strong use of quarantines. We stress that the primary objective of keeping 94% of the susceptible population infection free is maintained. Again, Fig. 5d demonstrates the higher importance of quarantine among students with respect to staff. This shows the predominant role played by student quarantine in controlling the epidemic.

**Fig. 5.**
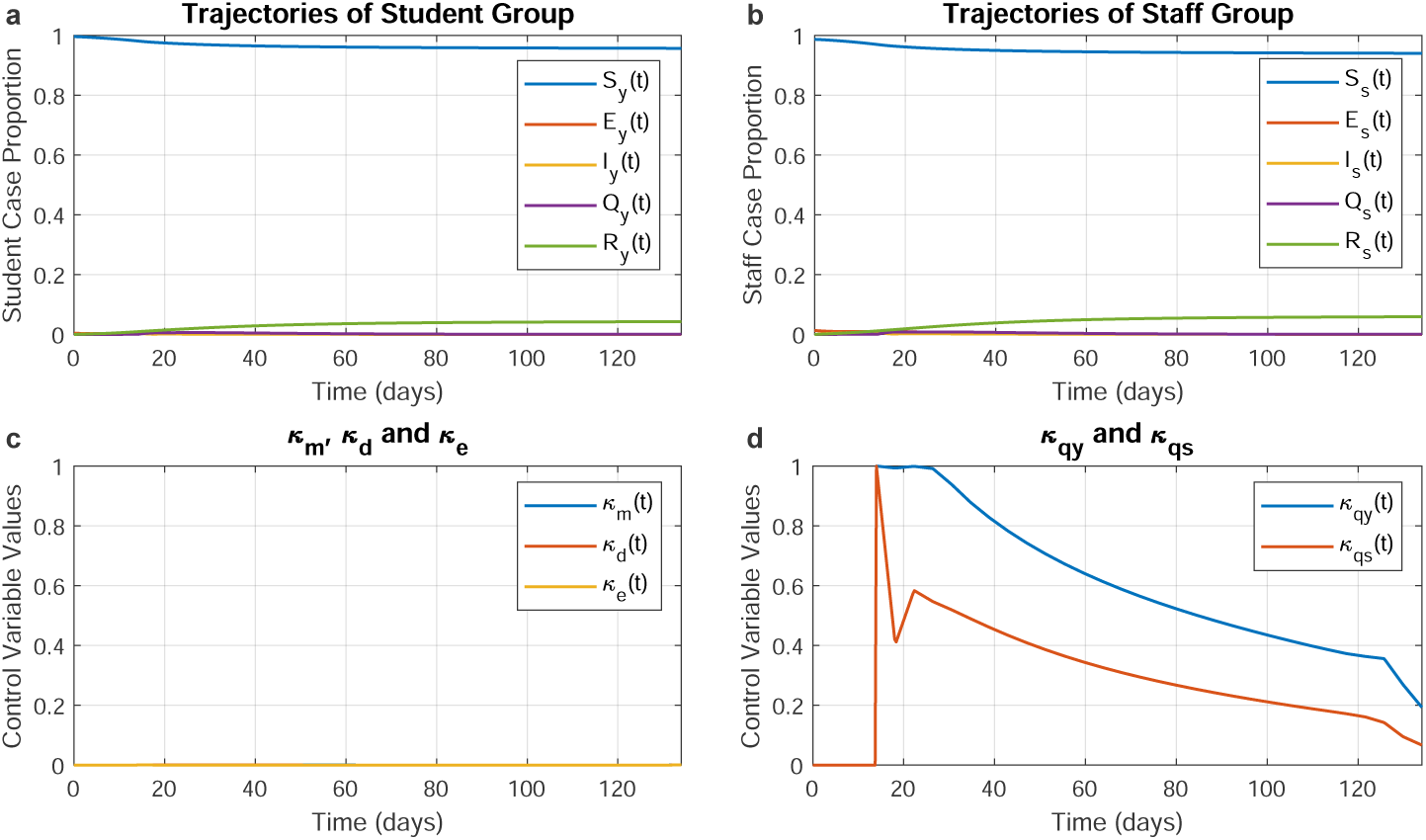
Optimal trajectories for minimum use of non-quarantine interventions. Optimal trajectories when the departments would like to minimise use of masks, social distancing and environmental disinfection. **a**,**b**, The epidemic evolution among students and staff, respectively. **c**, The optimal strategies for mask wearing (*κ*_*m*_), social distancing (*κ*_*d*_), and environmental disinfection (*κ*_*e*_). **d**, The optimal strategies for mandatory quarantine on infected students (*κ*_*qy*_) and infected staff (*κ*_*qs*_).

#### Minimum quarantine

In this scenario we want to minimise the use of quarantine, but we allow a strong use of mask wearing, social distancing and environmental disinfection. The resulting optimal trajectories are shown in Fig. 6. As a result, the quarantine control variables are zero and the other three interventions are major tools to resolve the epidemic in this scenario. Fig. 6c clearly shows the primary role of mask wearing and social distancing in keeping the infection under control. The figure also shows that environmental disinfection in comparison play little role when strong mask wearing and social distancing are in place.

**Fig. 6.**
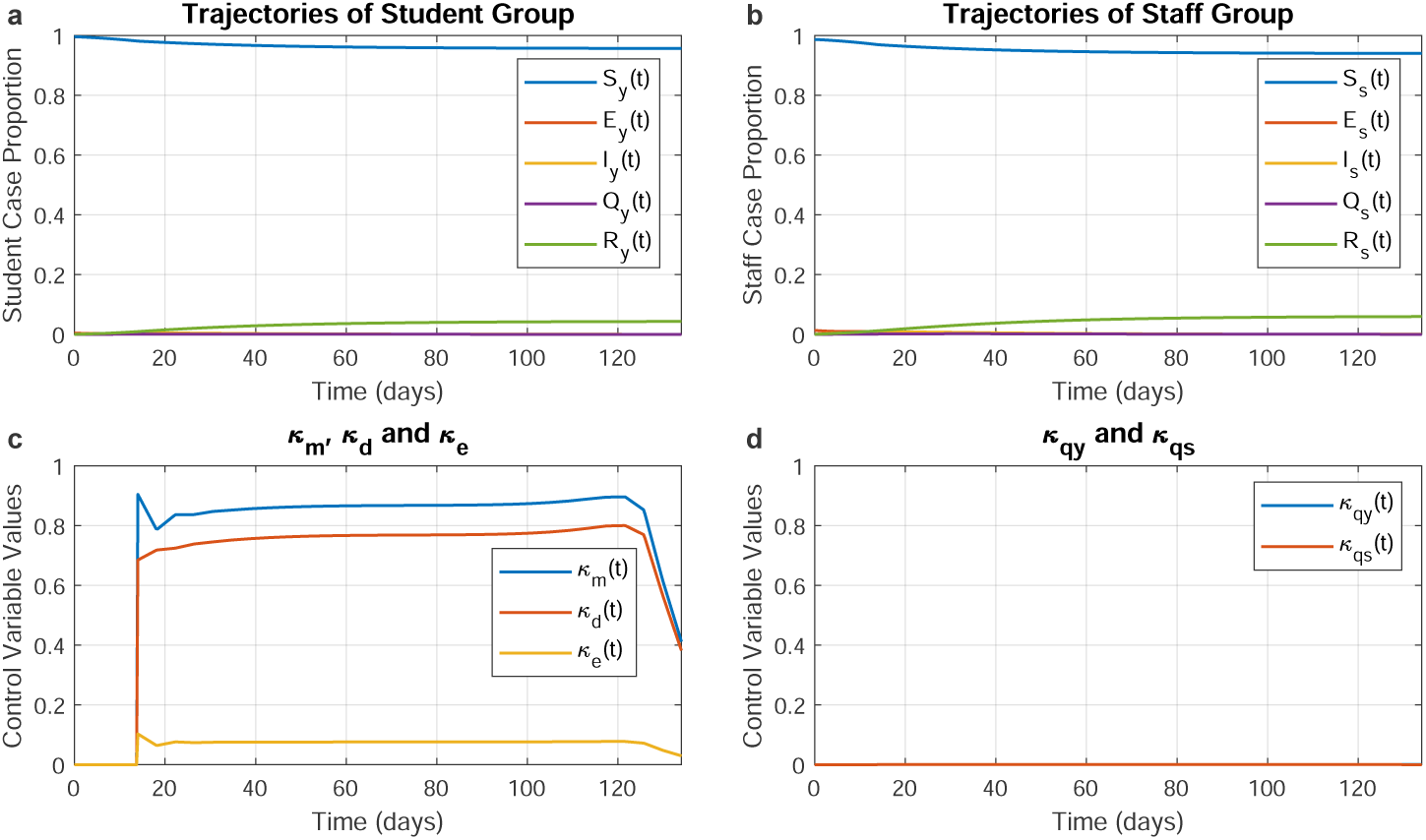
Optimal trajectories for the minimum quarantine scenario. Optimal trajectories when the department would like to minimise the enforcement of mandatory quarantines. **a**,**b**, The epidemic evolution among students and staff, respectively. **c**, The optimal strategies for mask wearing (*κ*_*m*_), social distancing (*κ*_*d*_), and environmental disinfection (*κ*_*e*_). **d**, The optimal strategies for mandatory quarantine on infected students (*κ*_*qy*_) and infected staff (*κ*_*qs*_).

## 2 Discussion

The figures show an emerging behaviour: non-pharmaceutical interventions have different importance and this importance arises mathematically from the evolution of the epidemic. In a typical university department composed of 1200 students and 150 staff, with a vaccination rate of 68% for students and 78.8% for staff (see Methods, [27]) we see that the implementation of non-pharmaceutical interventions is still fundamental to reduce the number of infections to one tenth of the number of infections appearing in a completely uncontrolled scenario.

The ranking that arises from the study is as follows: wearing masks is the most effective measure among the considered interventions. Keeping social distance is ranked close second. This priority of mask wearing is reasonable in a university, where close contact is often unavoidable. Furthermore, we can also see that environmental disinfection seems to be far less necessary if both measures are strongly enforced. As for the enforcement of mandatory quarantines, the result yields that quarantine of symptomatic students is more significant than quarantine of staff. This ranking is robust with respect to model parameters. This is shown in the sensitivity study presented in Figs. A1 to A8 in Appendix A.

From a practical perspective, the university should emphasize mask wearing and social distancing when on-campus teaching is resumed, especially among students. The study also suggests that the university should invest particular effort in identifying and quarantining infected students.

The proposed model and optimal control framework can be easily used to assess other scenarios, e.g. other objectives or other constraints. This tool can assist universities in predicting and managing the evolution of the epidemic. Moreover, our findings demonstrably reflect the importance of different non-pharmaceutical interventions and help to tackle the trade-off between high-quality teaching and limiting COVID-19 infections. This is crucial at a time in which universities are under pressure to increase on-campus activities.

Our study also shows a gap in the epidemiological research of COVID-19 regarding the evolution of the pandemic in small environments. There are plenty of small environments, such as schools and companies that have their very unique population structure (*i*.*e*. populations with different degrees of interaction, vaccination rates and serious symptoms) and require design tools to assess the best interventions to be implemented in order to maximise in-person activities while keeping the infections under control.

We also point out that some factors are not explicitly considered in our study. Firstly, the model does not consider the infections brought from outside the campus. We omitted this aspect because we wanted to focus on the study of the priority of different mitigation measures. Another limitation is that the input variables (the interventions) in our optimal control problem are continuous in magnitude. It may be difficult to give practical significance to the numerical values representing the interventions. However, we stress that the optimal trajectories are used here only to compare the relative importance of different interventions. Further research can be done on discretizing the magnitude of the input variables into specific levels that correspond to scientifically-defined interpretable practical meanings.

## 3 Methods

### 3.1 Uncontrolled University Model

The proposed double SEIQR model is described by 11 differential equations, 5 associated to the epidemic evolution of students, 5 associated to the epidemic evolution of staff, and 1 representing environmental infection. We first introduce, describe and analyse the uncontrolled model. In Section 3.3 we modify the model by introducing the control variables that represent non-pharmaceutical interventions. The double uncontrolled SEIQR model is described by

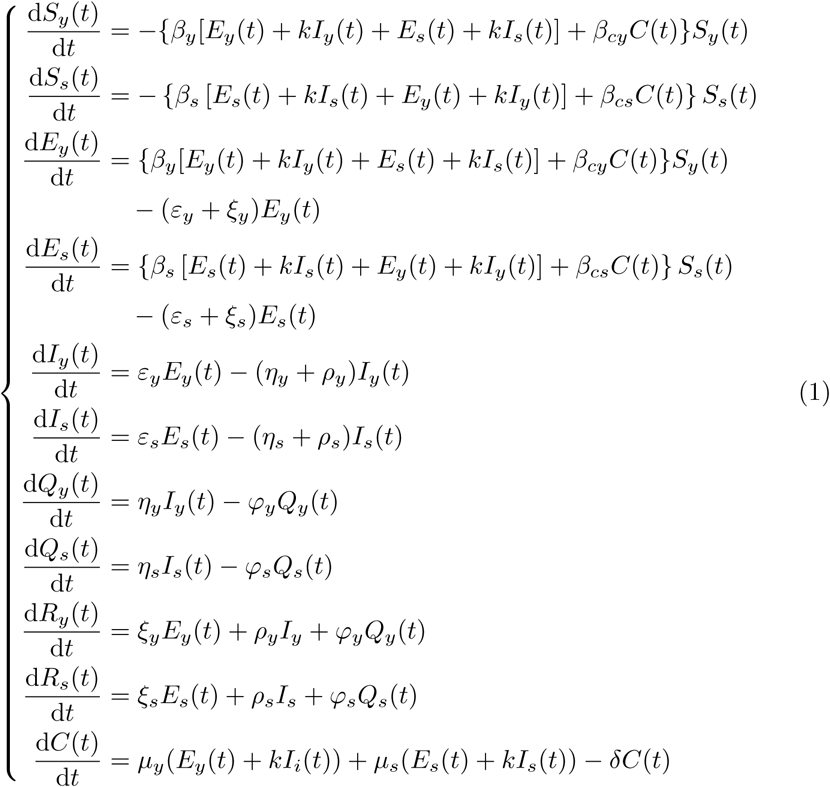

where all model parameters are denoted by Greek letters with specific biological meanings. We now provide a detailed explanation of each parameter. We stress that this model is uncontrolled, so the values discussed below are for the baseline scenario, *i*.*e*. no intervention is implemented. Also, the parameters are firstly introduced for unvaccinated population and then modified according to the UK vaccination proportions.

- Individual infection rates *β*_*y*_, *β*_*s*_ *kβ*_*y*_, *kβ*_*s*_ The individual infection rates represent the average number of susceptible individuals who can be infected by a virus carrier via direct contacts in unit time. *β*_*y*_ indicates student-to-student and staff-to-student infection rates. *β*_*s*_ indicates staff-to-staff and student-to-staff infection rates. In other words, we assume that student-to-student and staff-to-student rates are the same and staff-to-staff and student-to-staff rates are the same. *β*_*s*_ and *β*_*y*_ are the infection rates of the asymptomatic compartments. Considering the age difference, it is reasonable to expect that *β*_*y*_ *< β*_*s*_ because staff are more likely to get infected. *kβ*_*s*_ and *kβ*_*y*_ are the infection rates of the symptomatic group. Since sneezing and coughing play a major role in the direct transmission of the virus, the symptomatic carriers generally have larger infection rates, *i*.*e. k >* 1. Here it is assumed that *k* has the same value among students and staffs. Referring to the scenario 3 of pandemic planning produced by the CDC [30], *k* is generally 4. However, in this confined environment case, *k* should be smaller because asymptomatic subjects can spread the virus more easily. We selected a value of *k* = 1.5. According to the study conducted by Leontitsis et al. [22], the general infection rate *β* is 0.1466. This value is expected to be larger in a confined space because of the higher number of direct contacts between people. In summary, putting together all these data and observations, the parameters have been selected as *β*_*y*_ = 0.163, *β*_*s*_ = 0.225, *kβ*_*y*_ = 0.2445 and *kβ*_*s*_ = 0.3375.
- Environmental infection rates *β*_*cy*_, *β*_*cs*_ These parameters represent how many susceptible people are infected by the contaminated environment in unit time. They are properties of the virus in the environment. There is no clear value in the literature and we estimate the value of *β*_*cy*_ to be 0.171 (based on the expected basic reproduction number). Moreover, it is reasonable to expect that the ratio *β*_*s*_*/β*_*y*_ equals the ratio *p* = *β*_*cs*_*/β*_*cy*_. Then *β*_*cs*_ = *pβ*_*cy*_ = 0.236.
- Probability of becoming symptomatic *ε*_*y*_, *ε*_*s*_ They are the inverse of the average incubation period. According to [31], this average period is 5 days. Considering that staff are of higher age, we set *ε*_*y*_ = 1*/*5 = 0.2 and *ε*_*s*_ = 1*/*10 = 0.1.
- Probability of recovery from an asymptomatic state *ξ*_*y*_, *ξ*_*s*_ Similarly, the inverses of these quantities denote the average number of days spent by exposed/asymptomatic subjects to recover (*i*.*e*. they present no symptom during the whole period). Referring to [30], this portion accounts for 15%. Since *ε*_*y*_ = 0.2 and *ε*_*s*_ = 0.1 (corresponding to 85% of the population). Then *ξ*_*y*_ = 0.0353 and *ξ*_*s*_ = 0.0176.
- Isolation rates *η*_*y*_, *η*_*s*_ These parameters denote the proportion of symptomatic individuals who are isolated due to serious illness or mandatory quarantine. Since we initially model the unrestricted situation (i.e. no quarantines), infected individuals are at this point isolated mainly due to hospitalization. According to the survey conducted and reported by [32], the values are *η*_*y*_ = 0.06 and *η*_*s*_ = 0.106.
- Recovery rates of infected individuals *ρ*_*y*_, *ρ*_*s*_ 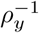 and 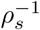 indicate the average time for infected people which are not isolated to recover. If mandatory quarantine is not implemented, mild cases will not isolate. These two parameters mainly describe the recovery rate of this group. According to [33], the average length of recovery is approximately 14 days. In mild cases, we set this length at 10 days for students, and 18 days for staff. Thus, *ρ*_*y*_ = 1*/*10 and *ρ*_*s*_ = 1*/*18.
- Recovery rates of quarantined individuals *ϕ*_*y*_, *ϕ*_*s*_ 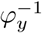 and 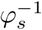 indicate the average time for “quarantined” individuals to recover. In the baseline scenario this refers to hospitalised individuals. It may take six-nine weeks for severe cases to recover [34]. We set *ϕ*_*y*_ = 1*/*40 while *ϕ*_*s*_ = 1*/*55.
- Virus shedding rates to the environment *µ*_*y*_, *µ*_*s*_, *kµ*_*y*_, *kµ*_*s*_ These parameters measure the spread of the virus from asymptomatic/symptomatic individuals to the environment, with the effects brought by symptomatic subjects being higher. Similarly to the case of *β*_*cy*_ and *β*_*cs*_, there is no clear value in the literature for these parameters. In this “small environment” model we expect the values to be *µ*_*y*_ = *µ*_*s*_ = *µ* = 0.25 (based on the expected basic reproduction number).
- Virus decaying rate in the environment *δ* This measures the speed of decay of the virus in the small environment. Since the airborne virus could stay in aerosol for up to 1 day and survive on the surface for longer [35–37], this rate *δ* is set at 0.7.

We denote *N*_*t*_ to be the total population in the department. The total number of students is represented by *N*_*y*_ and number of staff is labelled by *N*_*s*_. A summary of the meaning the parameters is given in Table 1.

**Table 1.** Model Parameter Definitions.

| Parameters | Definitions |
| --- | --- |
| $\beta_y, \beta_s$ | Infection rate for asymptomatic students/staff to susceptible students/staff (including cross infections) |
| $k\beta_y, k\beta_s$ | Infection rate for symptomatic students/staff to susceptible students/staff (including cross infections) |
| $\beta_{cy}, \beta_{cs}$ | Infection rate from uncleaned environment to susceptible students/staff |
| $\varepsilon_y, \varepsilon_s$ | Probability that an asymptomatic student/staff becomes symptomatic |
| $\xi_y, \xi_s$ | Probability that an asymptomatic student/staff recovers without symptoms |
| $\eta_y, \eta_s$ | Isolation rate of symptomatic student/staffs |
| $\rho_y, \rho_s$ | Recovery rate of infected student/staffs |
| $\varphi_y, \varphi_s$ | Recovery rate of quarantined students/staffs |
| $\mu_y, \mu_s$ | Environmental shedding rate by asymptomatic students/staffs |
| $k\mu_y, k\mu_s$ | Environmental shedding rate by symptomatic students/staffs |
| $\delta$ | Decaying rate of virus in the environment |

These initial values refer to studies on the COVID-19 epidemic before vaccination. We now describe how the parameters are adapted to a vaccinated population. According to the UK data provided by [27], about 68% of young adults between 18 and 24 years old have been vaccinated by the 1^*st*^ November 2021. This quantity become 78.7% among people aged between 25 and 64. Since vaccines utilized in the UK can reduce COVID-19 infections by around 65% [28, 29], we can see that the 68% vaccinated students will have 65% less probability of getting infected. The same happens to the 78.7% of staffs. Therefore, the average reductions in *β*_*y*_ and *β*_*cy*_ are 0.68 *×* (1 *−* 0.65) + 0.32 = 0.558. The average reductions in *β*_*s*_ and *β*_*cs*_ are 0.787 × (1−0.65)+0.213 = 0.488. Consequently, due to vaccinations in the current situation, infection rates in this university model with mixed vaccinated/unvaccinated population becomes: *β*_*y*_ = 0.0910, *β*_*cy*_ = 0.0954, *β*_*s*_ = 0.1098, and *β*_*cs*_ = 0.1152. Since vaccines can also reduce the probability of symptomatic infections and of severe illness [38], other parameter values are also tuned accordingly. This adjustment changed the *R*_0_ from 2.50 (totally unvaccinated and no interventions) to 1.40 (mixed population but still no intervention). Since it is still greater than 1, the COVID-19 epidemic will still develop in the university model if no other control or prevention measures are introduced. The values of the parameters of the model before and after vaccinations are listed in Table 2.

**Table 2.** Model Parameter Values.

| Parameters | Before Vaccination | After Vaccination |
| --- | --- | --- |
| $\beta_y$ | 0.163 | 0.0910 |
| $\beta_s$ | 0.225 | 0.1098 |
| $k$ | 1.5 | 1.5 |
| $\beta_{cy}$ | 0.171 | 0.0954 |
| $\beta_{cs}$ | 0.236 | 0.1152 |
| $\varepsilon_y$ | 0.2 | 0.2 |
| $\varepsilon_s$ | 0.1 | 0.1 |
| $\xi_y$ | 0.0353 | 0.0857 |
| $\xi_s$ | 0.0176 | 0.0429 |
| $\eta_y$ | 0.06 | 0.012 |
| $\eta_s$ | 0.106 | 0.0212 |
| $\rho_y$ | 0.1 | 0.125 |
| $\rho_s$ | 0.0556 | 0.0833 |
| $\varphi_y$ | 0.025 | 0.0714 |
| $\varphi_s$ | 0.0182 | 0.0714 |
| $\mu_y$ | 0.25 | 0.25 |
| $\mu_s$ | 0.25 | 0.25 |
| $\delta$ | 0.7 | 0.7 |

### 3.2 Analysis of the Uncontrolled University Model

#### 3.2.1 Equilibrium Points

Denote *x* = [*S*_*y*_, *S*_*s*_, *E*_*y*_, *E*_*s*_, *I*_*y*_, *I*_*s*_, *Q*_*y*_, *Q*_*s*_, *R*_*y*_, *R*_*s*_, *C*]^*T*^. By equating all derivatives in (1) to zero, it is easy to determine the equilibria 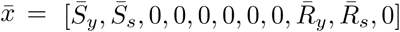, where

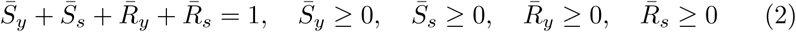

These equilibria imply that at the end of pandemic, the individuals are either susceptible to or recovered from the disease.

#### 3.2.2 Basic Reproduction Number

The basic reproduction number is a crucial criterion to measure the average number of susceptible people that could potentially be infected by a primary case [39]. This parameter is highly dependent on the fraction of the susceptible population and it provides information about the potential of the epidemic outbreak. If *R*_0_ *<* 1 the disease will gradually disappear. If *R*_0_ *>* 1 increasingly more people will be infected.

Derivation of the basic reproduction number for the uncontrolled university model is based on the next generation matrix method described by [40–42]. The university model (1) has five infectious compartments: *E*_*y*_, *E*_*s*_, *I*_*y*_, *I*_*s*_ and *C*. We collect these in the infection state *x*_*if*_ = [*E*_*y*_, *E*_*s*_, *I*_*y*_, *I*_*s*_, *C*]^*T*^. Let *F* denote the rate of increase of secondary cases and *V* denote the progression rate. Accordingly, *x*_*if*_ obeys the equation

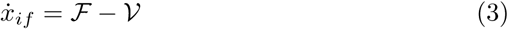

where *ℱ* and *𝒱* given by

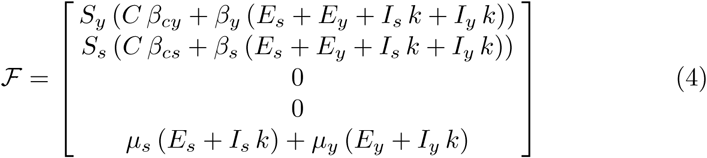

and

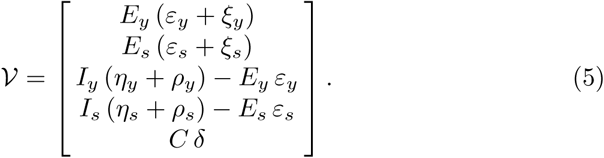

We linearise equation (3) around the equilibrium and we denote the Jacobians of *ℱ* and *𝒱* as *ℱ* and *𝒱*, respectively. Thus, we obtain

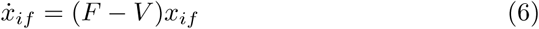

where

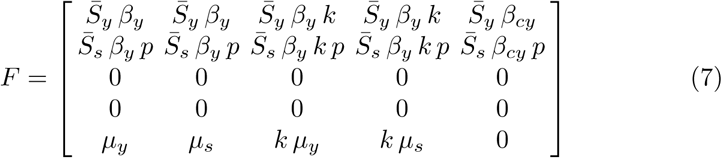

and

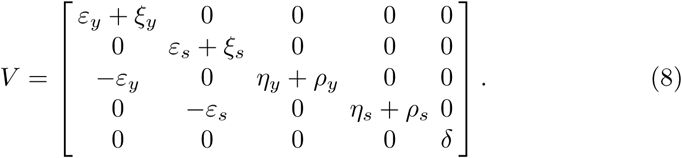

Denoting *σ*_*y*_ = *η*_*y*_ + *ρ*_*y*_ and *σ*_*s*_ = *η*_*s*_ + *ρ*_*s*_, the next generation matrix *K* is therefore derived as

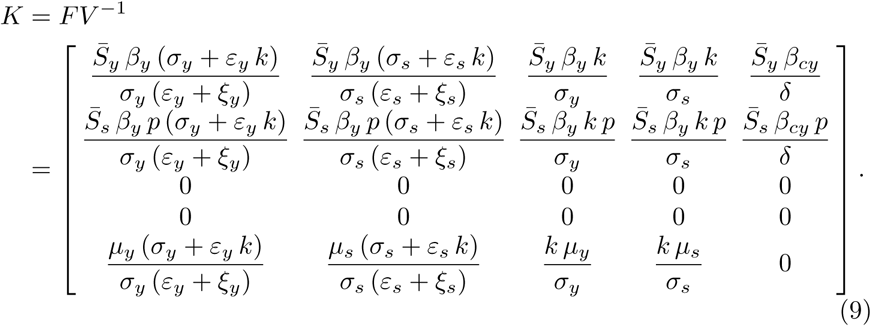

The obtained matrix *K* is nonnegative and has rank 2. In particular, it has three zero eigenvalues and two positive eigenvalues. According to [40], *R*_0_ is the spectral radius of *K, i*.*e*. its largest eigenvalue. By computing det(*λI − K*), the characteristic polynomial is

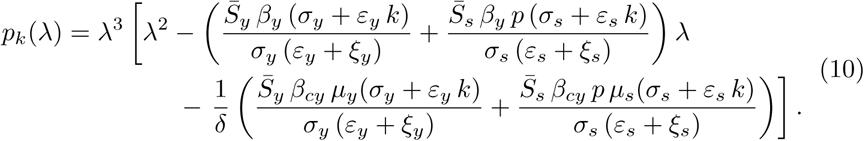

We recall that we have assumed that *µ*_*y*_ = *µ*_*s*_ = *µ*, which means that students and staff have equal rates of spreading the virus into the environment. The two non-zero eigenvalues can be derived by finding the roots of the polynomial

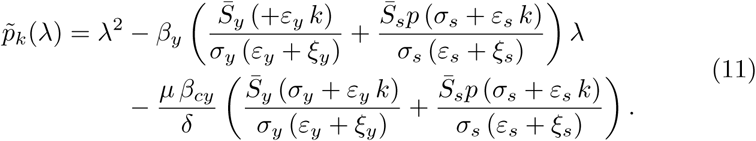

Denote

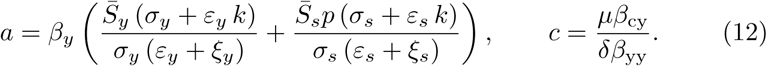

Then

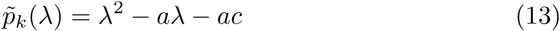

Since the constants *a* and *c* are both positive, the quadratic equation has two real roots and *R*_0_ will be the larger one. Therefore, we can express *R*_0_ as

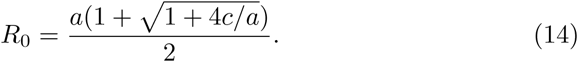

In the next section we show how *R*_0_ is related to the stability of the equilibrium point.

#### 3.2.3 Stability Analysis

##### Proposition 1

*If R*_0_ *<* 1, *the equilibrium points* 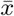 *of the uncontrolled university model (1) is asymptotically stable*.

*Proof* Model (1) can be reformulated into a feedback interconnection. Compartments *E*_*y*_, *E*_*s*_, *I*_*y*_, *I*_*s*_, *Q*_*y*_, *Q*_*s*_, *C* form a positive linear subsystem with output feedback topology. Defining *x*_*l*_ = [*E*_*y*_, *E*_*s*_, *I*_*y*_, *I*_*s*_, *Q*_*y*_, *Q*_*s*_, *C*^*T*^, *y*_*s*_ = [*S*_*y*_, *S*_*s*_]^*T*^, *y*_*R*_ = [*R*_*y*_, *R*_*s*_]^*T*^, the subsystem can be formulated as

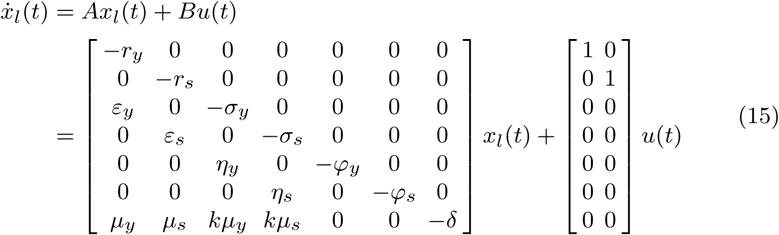

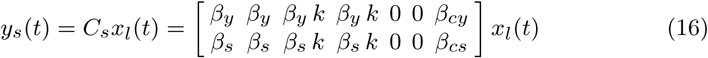

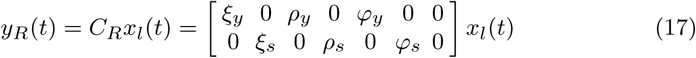

where *r*_*y*_ = *ε*_*y*_ + *η*_*y*_, *r*_*s*_ = *ε*_*s*_ + *η*_*s*_. Since output *y*_*R*_ does not contribute to the variations of the state variable *x*_*l*_, we can represent the overall system dynamics by the output feedback loop as

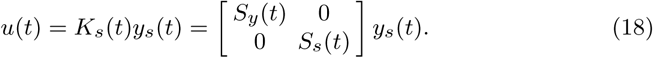

Derivatives of the remaining compartments can be calculated as

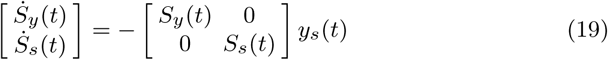

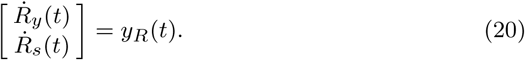

Note that the system has a time-varying feedback *K*_*s*_(*t*). Around the equilibrium 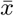, the system behaviour is determined by using he constant feedback term 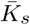 as

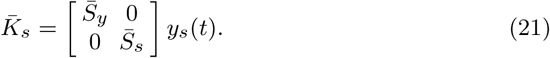

Therefore, the dynamics of the original model is equivalent to that of this closed-loop system. To study its stability, we firstly derive the closed-loop system matrix *A*_*cl*_ as

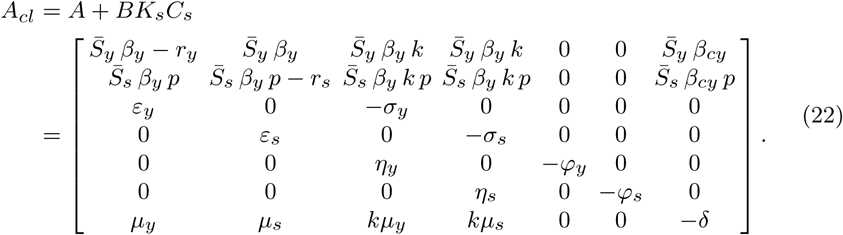

To determine its closed-loop poles, we compute its characteristic equation, *i*.*e*. det(*λI−A*_*cl*_). Since all parameters are positive, *A*_*cl*_ must have two negative eigenvalues at *−φ*_*y*_ and *−φ;*_*s*_. The remaining five eigenvalues are the roots of the polynomial *p*_5_(*λ*):

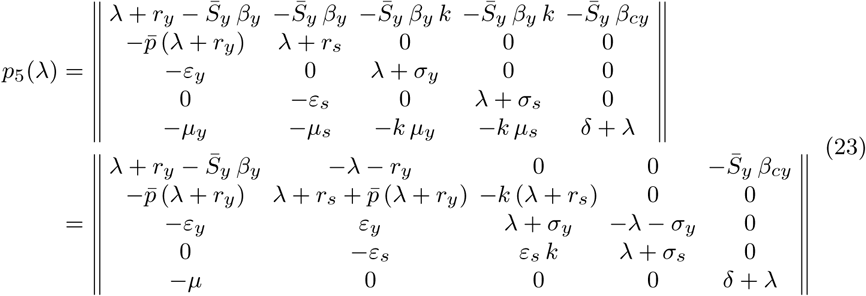

where we have defined 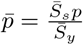 and used that *µ*_*y*_ = *µ*_*s*_ = *µ* according to the previous analysis. The polynomial is finally derived as

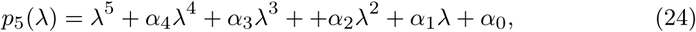

where

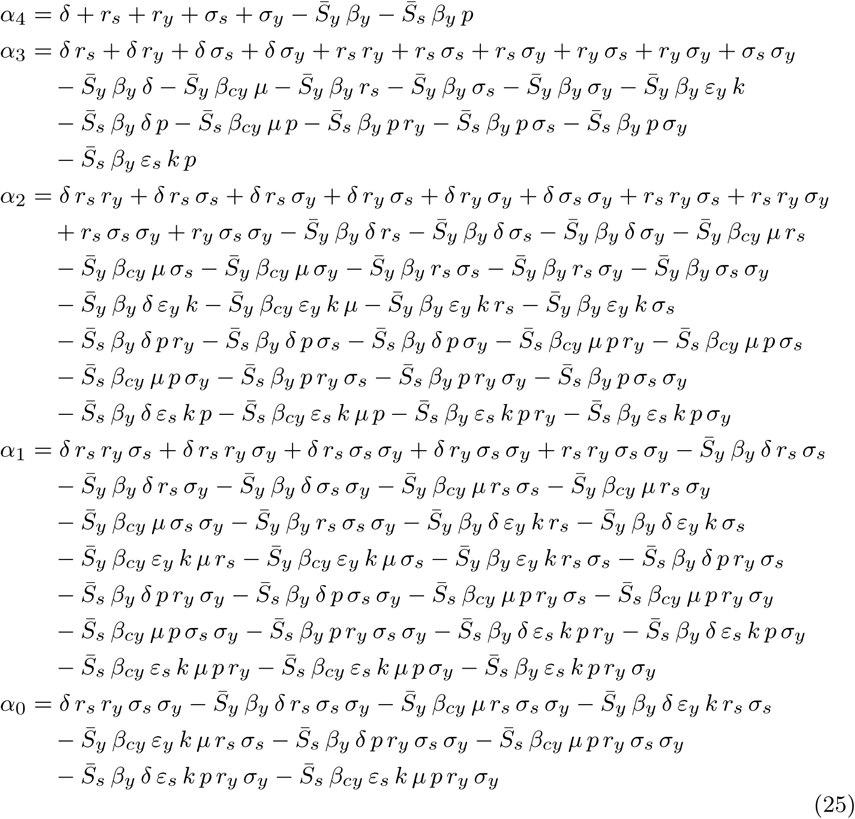

The conditions to obtain a stable equilibrium point can be determined using the Routh-Hurwitz stability criterion. The Routh table is given in Table 3.

**Table 3.** Routh Table.

|  |  |  |  |
| --- | --- | --- | --- |
| $\lambda^5$ | 1 | $\alpha_3$ | $\alpha_1$ |
| $\lambda^4$ | $\alpha_4$ | $\alpha_2$ | $\alpha_0$ |
| $\lambda^3$ | $b_{31} = -\frac{1}{a_4}(\alpha_2 - \alpha_4 \alpha_3)$ | $b_{32} = -\frac{1}{a_4}(\alpha_0 - \alpha_1 \alpha_4)$ | 0 |
| $\lambda^2$ | $b_{21} = -\frac{1}{b_{31}}(\alpha_4 b_{32} - \alpha_2 b_{31})$ | $\alpha_0$ | 0 |
| $\lambda^1$ | $b_{11} = -\frac{1}{b_{21}}(\alpha_0 b_{31} - b_{32} b_{21})$ | 0 | 0 |
| $\lambda^0$ | $\alpha_0$ | 0 | 0 |

From the Routh-Hurwitz stability criterion follows that the equilibrium point is asymptotically stable if and only if *α*_4_ *>* 0, *b*_31_ *>* 0, *b*_21_ *>* 0, *b*_11_ *>* 0 and *α*_0_ *>* 0. We evaluate these coefficients numerically for different values of *R*_0_. The results are shown in Table 4 for *R*_0_ *<* 1 and in Table 5 for *R*_0_ *>* 1. Since *α*_0_ *>* 0 in Table 4 and *α*_0_ *<* 0 in Table 5, the Routh-Hurwitz stability criterion confirms that the *R*_0_ found in (14) is consistent with the expected epidemic dynamics.

**Table 4.**
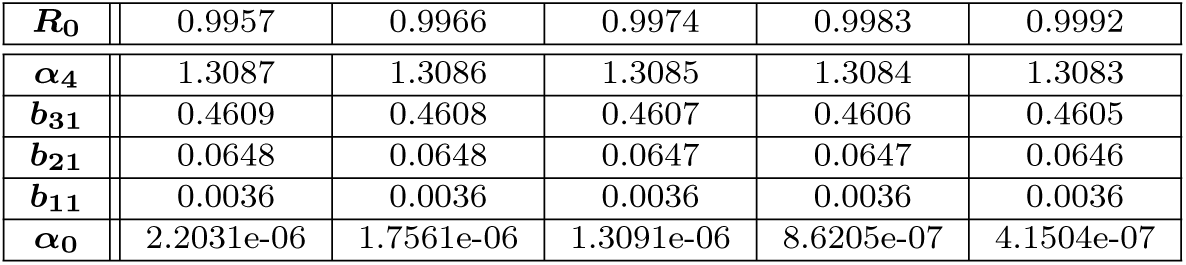
Values of the first column of the Routh table when *R*_0_ *<* 1.

**Table 5.** Values of the first column of the Routh table when *R*_0_ *>* 1.

|  |  |  |  |  |  |
| --- | --- | --- | --- | --- | --- |
| $R_0$ | 1.0001 | 1.0009 | 1.0018 | 1.0027 | 1.0036 |
| $\alpha_4$ | 1.3082 | 1.3081 | 1.3079 | 1.3078 | 1.3077 |
| $b_{31}$ | 0.4604 | 0.4603 | 0.4602 | 0.4601 | 0.4600 |
| $b_{21}$ | 0.0646 | 0.0646 | 0.0646 | 0.0645 | 0.0645 |
| $b_{11}$ | 0.0036 | 0.0036 | 0.0036 | 0.0036 | 0.0036 |
| $\alpha_0$ | -3.1972e-08 | -4.7898e-07 | -9.2600e-07 | -1.3730e-06 | -1.8200e-06 |

### 3.3 Control of the University Model

#### 3.3.1 Formulation of the Controlled University Model

In this study we consider five non-pharmaceutical interventions that the university can implement.

1. Compulsory mask wearing: how strongly this measure is implemented is represented by the normalised variable 0 *≤ κ*_*m*_ *≤* 1.
2. Keep safe social distance: how strongly this measure is implemented is represented by the normalised variable 0 *≤ κ*_*d*_ *≤* 1.
3. Environment disinfection: how strongly this measure is implemented is represented by the normalised variable 0 *≤ κ*_*e*_ *≤* 1.
4. Mandatory quarantines: how strongly these measures are implemented is represented by the normalised variables 0 *≤ κ*_*qy*_ *≤* 1 (for students) and 0 *≤ κ*_*qs*_ *≤* 1 (for staff).

A group of five variables is initially defined to represent the reduction factors in the infection rates, shedding rates, environmental decaying rate, and isolation rates. Denoting these factors as *u* = [*u*_*p*_, *u*_*m*_, *u*_*e*_, *u*_*qy*_, *u*_*qs*_]^*T*^, the university model at this stage is expressed as

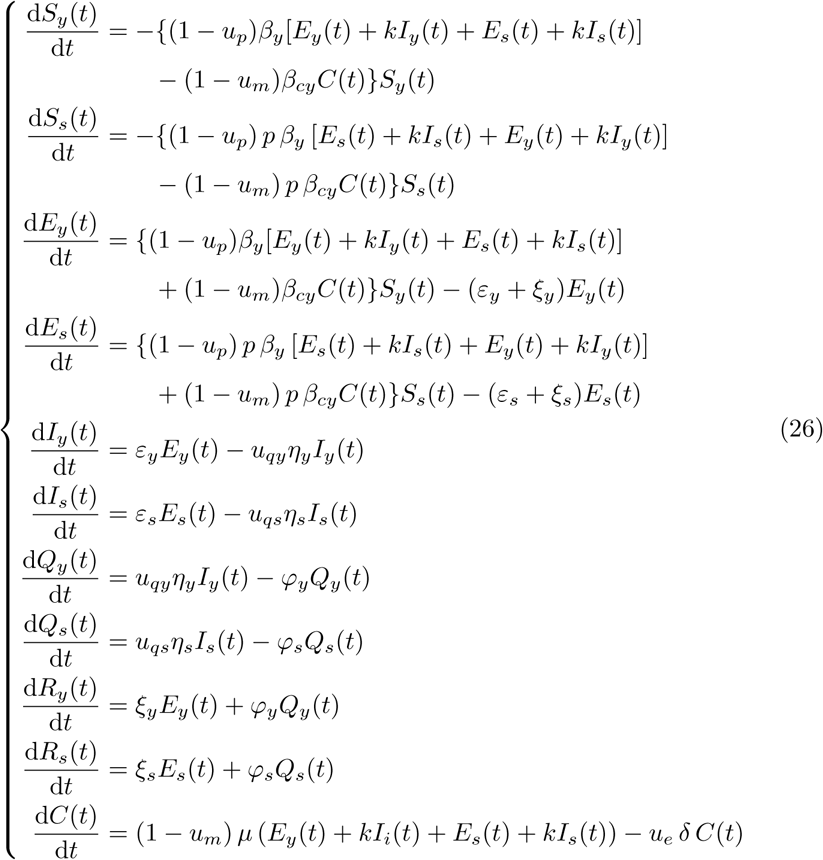

Note that *u*_*p*_, *u*_*m*_, *u*_*e*_, *u*_*qy*_*η*_*y*_, and *u*_*qs*_*η*_*s*_ should vary within [0, 1].

Since a reduction factor can be influenced by multiple interventions, we need to identify the relationship between the reduction factors (*u*’s) and intervention variables (*κ*’s).

1. Reduction of interpersonal infection rates *β*_*y*_ and *β*_*s*_ The person-to-person infection rates are directly influenced by two measures: wearing masks and keeping social distance. Wearing masks could reduce the probability of infections during contacts while social distancing could reduce the number of direct contacts between individuals. Study conducted by Karaivanov et al, [43] argued that the mandatory mask-wearing policy in confined spaces could reduce the number of infected cases by up to 40% weekly. Furthermore, Jarvis et al. [44] conducted a survey which showed that the physical distancing could reduce the number of direct contacts by 74%. However, since this survey might have selection and recall bias, the actual result should be lower than 74% to obtain a conservative estimate. In this case, we set the maximum reduction at 65%. Hence, 1 *− u*_*p*_ = (1 *−* 0.4*κ*_*m*_)(1 *−* 0.65*κ*_*m*_).
2. Reduction of shedding rates and environment-to-person infection rates: *µ*_*y*_, *µ*_*s*_, *β*_*cy*_, *β*_*cs*_ Shedding rates as well as infections due to unclean environment can be reduced by the use of masks. According to laboratory-based investigations by [45, 46], masks could block approximately 50% to 70% droplets and aerosol, greatly reducing the transmission of virus. In this case, we set the maximum reduction of shedding rate to be 60% for a conservative estimate. Therefore, 1 *− u*_*m*_ = 1 *−* 0.6*κ*_*m*_.
3. Enhancement of virus environmental decaying rate *δ* The environmental decaying rate could be magnified by disinfection of the confined space. Recall that the uncontrolled decaying rate was *δ* = 0.7. It is hard to determine how much this rate is increased. A conservative estimate is that the rate is increased at the maximum by 30%, yielding a maximum new rate *u*_*e*_*δ* = 0.91. The linear relationship can be finally defined as *u*_*e*_ = 1 + 0.3*κ*_*e*_. While the analysis above is derived from considerations extracted from the literature, these still assumed an ideal enforcement of the interventions. Since the university cannot guarantee full compliance, we limit *κ*_*m*_, *κ*_*d*_, and *κ*_*e*_ to 70% of their values.
4. Quarantine enhancement Recall that in model (1) the quarantined populations were simply equivalent to the populations who developed serious symptoms and their rate were *η*_*y*_ and *η*_*s*_. To consider the effects of mandatory quarantines we replace *η*_*y*_ and *η*_*s*_ by *u*_*qy*_*η*_*y*_ and *u*_*qs*_*η*_*s*_, respectively. (*u*_*qy*_*η*_*y*_)^*−*1^ and (*u*_*qs*_*η*_*s*_)^*−*1^ now indicate the average time that unisolated symptomatic students/staff stay in campus before being detected by the university. We set these values to 2, meaning that the university takes two days in average to detect and isolate infected individuals after their symptoms develop. Thus, the maximum values of (*u*_*qs*_*η*_*s*_)^*−*1^ and (*u*_*qs*_*η*_*s*_)^*−*1^ are both 0.5. These values correspond to the situation where the enforcement of mandatory quarantines reaches the strongest degree, which means that *κ*_*qy*_ and *κ*_*qs*_ are 1. On the contrary, when mandatory quarantines are not implemented, i.e. *κ*_*qy*_ = *κ*_*qs*_ = 0, we want that the resulting quarantine rates *u*_*qy*_*η*_*y*_ and *u*_*qs*_*η*_*s*_ are still *η*_*y*_ and *η*_*s*_, respectively. These relations are formulated as *u*_*qy*_*η*_*y*_ = (0.5 *− η*_*y*_)*κ*_*qy*_ + *η*_*y*_ and *u*_*qs*_*η*_*s*_ = (0.5 *− η*_*s*_)*κ*_*qs*_ + *η*_*s*_.

In summary, the following relations hold

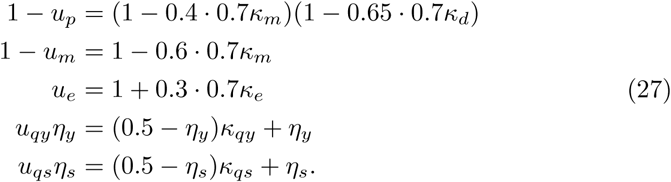

Substituting these relationships into equation (26), we obtain the final controlled university model

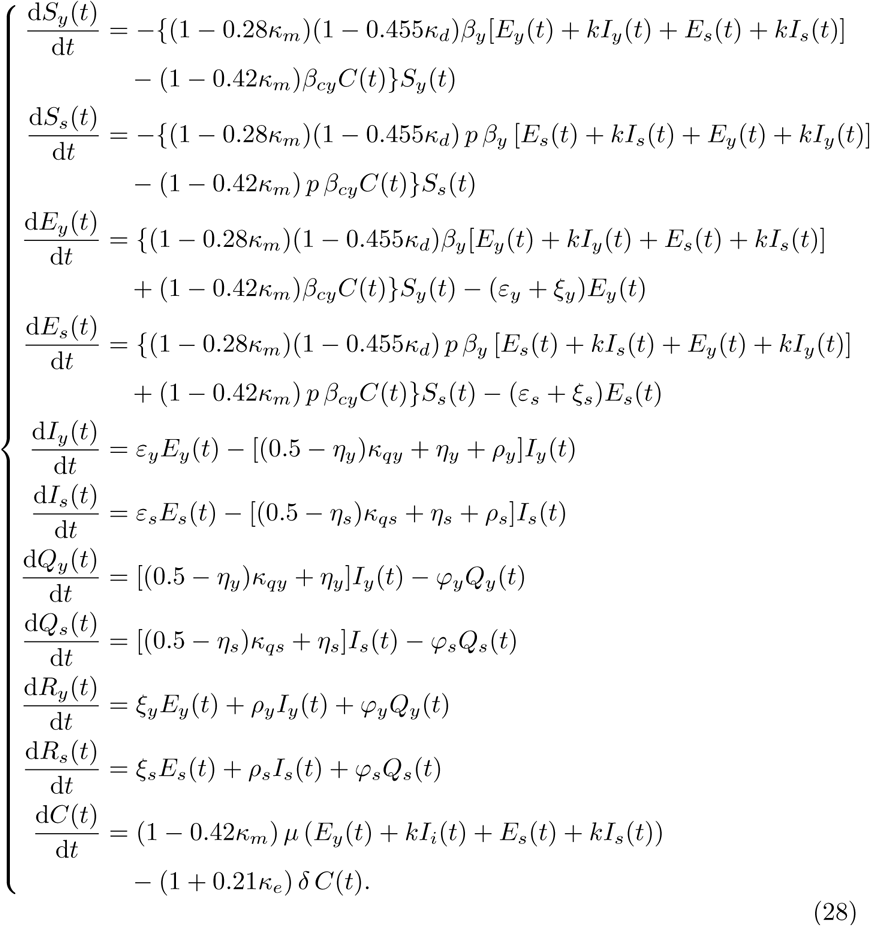

In this model, the variables *κ* = [*κ*_*m*_, *κ*_*d*_, *κ*_*e*_, *κ*_*qy*_, *κ*_*qs*_]^*T*^ are the control variables that need to be optimised.

#### 3.3.2 Formulation of the Optimal Control Problem

Now we formulate the optimal control problem that, once solved, provides the optimal trajectories that give the best combination of interventions to contain the epidemic within the university. The optimal control problem is defined as

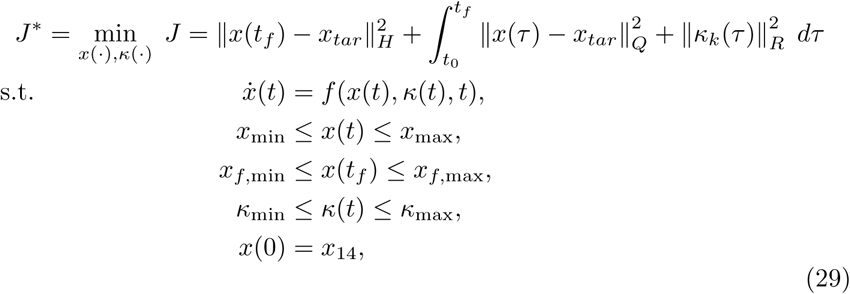

where *x*_*tar*_ represents the target state vector, _*H/Q/R*_ indicates the Euclidean matrix norm weighted by *H/Q/R* and the square matrices *H, Q* and *R* contain the weights for the final states, running states and running control variables, respectively. The values of *x*_*tar*_ and the weights are changed depending on the scenario that we need to solve (see next section). The problem has also constraints on both states and control variables. In fact, we need that the states lie between 0 and 1. Thus, even without further requirement the state constraints are at least *x*_min_ = *x*_*f*,min_ = [0, 0, 0, 0, 0, 0, 0, 0, 0, 0, 0]^*T*^ and *x*_max_ = *x*_*f*,max_ = [1, 1, 1, 1, 1, 1, 1, 1, 1, 1, 1]^*T*^. Similarly, the lower bounds of the control variables should be *κ*_min_ = [0, 0, 0, 0, 0]^*T*^ while their upper constrains are *κ*_max_ = [1, 1, 1, 1, 1]^*T*^. The initial condition sets the starting point of the system. We assume that the university starts to react two weeks after the first exposed subjects appear among students or staff. Thus, we first run the simulation of the original un-controlled model for 14 days and get the resulting state values on day 14, namely *x*_14_. Then this state is used as the initial condition for the optimisation problem, namely *x*(0) = *x*_14_. We finally require that the epidemic is eliminated within 120 days, so *t*_0_ = 0 and *t*_*f*_ = 120.

#### 3.3.3 Implementation of Four Different Scenarios

Balancing the trade-off between controlling the spread of COVID-19 and resuming the normal campus activity is the main question considered in this study. According to different trade-off’s between these two objectives, four scenarios are studied: minimum number of cases, minimum intervention, minimum non-quarantine interventions, and minimum quarantine interventions. Each scenario corresponds to different weight matrices and different path constraints.

For the first scenario (minimum number of cases), the university does not impose any limitations on the strength of the interventions to lead to the fastest mitigation of the epidemic. Mathematically, the controller is designed to maximise the number of susceptible people during the whole period, i.e., the proportions of the susceptible students/staff are maintained as close to one as possible. In contrast, the values of the other compartments need to be as close as possible to zero in this scenario. Therefore, we set the target state as

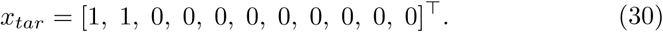

Meanwhile, the weight matrices are set as

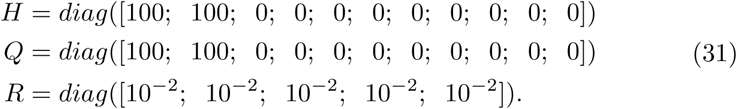

where in the matrix *R* we use 10^*−*2^ instead of 0 to improve numerical stability of the solver. The constraints on both states and control variables remain the same as the basic requirements discussed before, namely

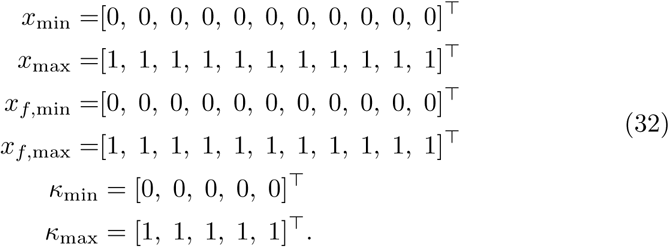

For the other three scenarios, we require that the number of susceptible students and staff cannot go below 94% of the total population (i.e. the number of infected individuals is equal or below 6%). We can easily achieve this by setting more restrictive state constraints. Consequently, we do not need to use the weights *H* and *Q* (also because it is difficult to intuitively understand the meaning of the weights on the states in these scenarios). Hence, we select *H* = 0 and *Q* = 0 and *x*_*tar*_ is not used. Additionally, to ensure that the epidemic is completely concluded after 134 days, the number of exposed and infected individuals should become zero at the final state. Recall that *N*_*y*_, *N*_*s*_, and *N*_*t*_ indicate the total number of student, staff, and the total population, respectively. Then the constraints on states should be

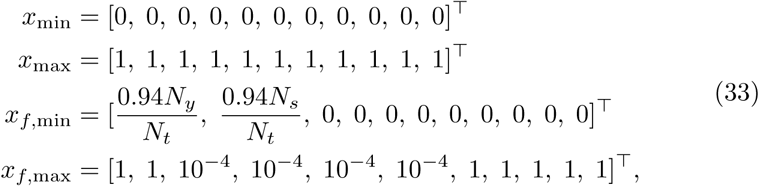

while the input constraints remain unchanged

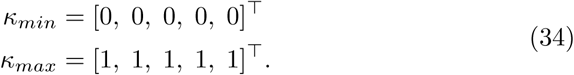

The only difference in the formulation of the other three scenarios is the value of the weight matrix *R*. In the minimum intervention scenario, the solver aims to minimize the total norm of the control signal while satisfying the 94% constraint. Thus, all control variables are heavily weighted and the matrix *R* is selected as

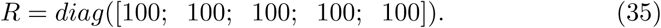

In the scenario of minimisation of non-quarantine interventions, the strength of first three interventions (wearing masks *κ*_*m*_, keeping social distance *κ*_*d*_ and disinfecting the environment *κ*_*e*_) are minimised, while the quarantines are not. In this situation, the matrix *R* becomes

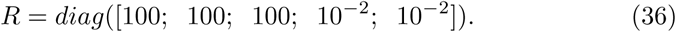

In the minimum quarantine scenario, the norm of quarantine rates for the infected students (*κ*_*qy*_) and staff (*κ*_*qs*_) are minimised. Since mandatory quarantine has the largest impact on campus activities, this scenario aims to find how the spread can be minimised while avoiding quarantine enforcement. Hence, the matrix *R* is

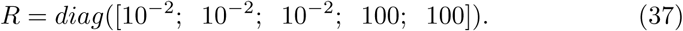

## Data Availability

All data produced in the present work are contained or referenced in the manuscript

## Code availability

The code is available at https://github.com/ZiruiNiu/University_Epidemic_Model_with_Control.git.

## Data availability

All model’s parameters taken from the literature are opportunely referenced in the main text. No further data is used.

## Appendix A Sensitivity Figures

**Fig. A1.**
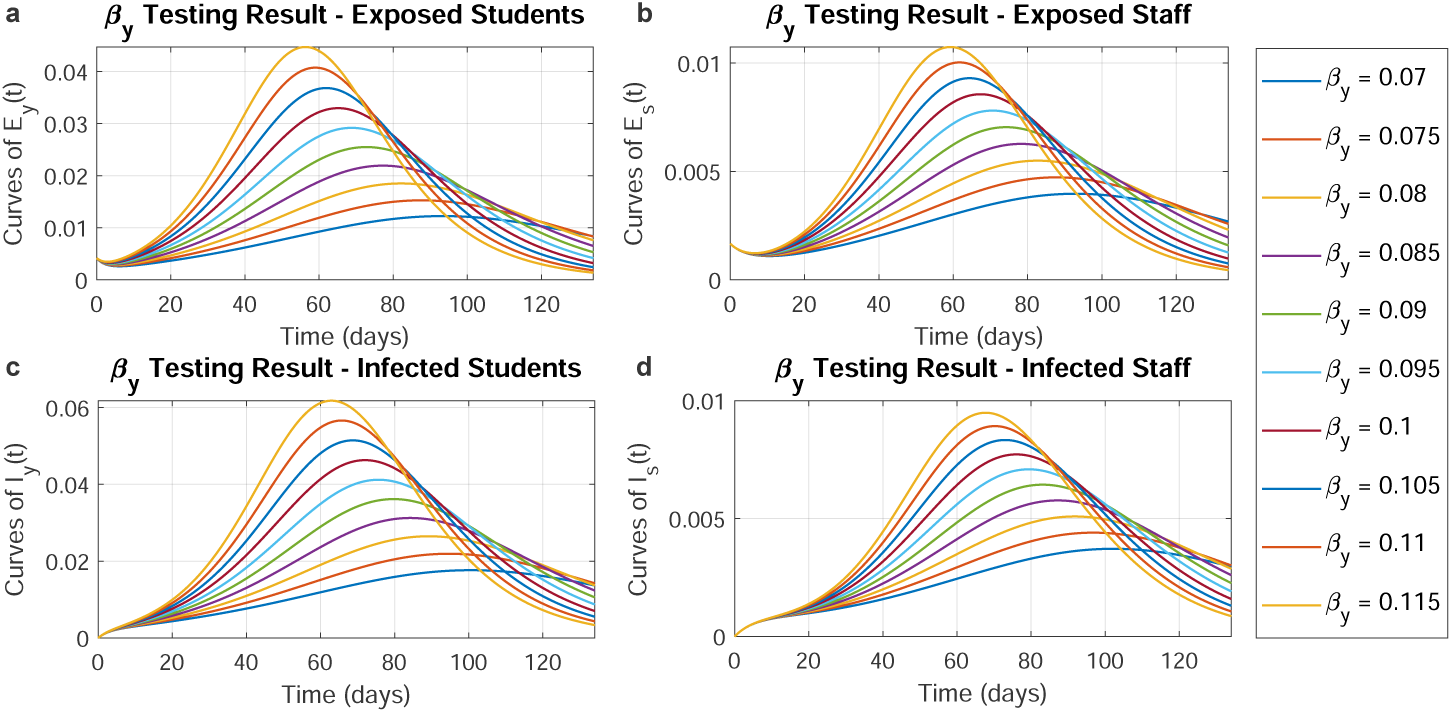
Numerical testing of student-to-student infection rate *β*_*y*_. Trajectories of cases among students and staffs when *β*_*y*_ varying from 0.07 to 0.115. With different *β*_*y*_, panels **a** and **b** show the proportion trajectories of exposed students and staffs respectively while panels **c** and **d** depict the proportion trajectories of infected students and staffs respectively.

**Fig. A2.**
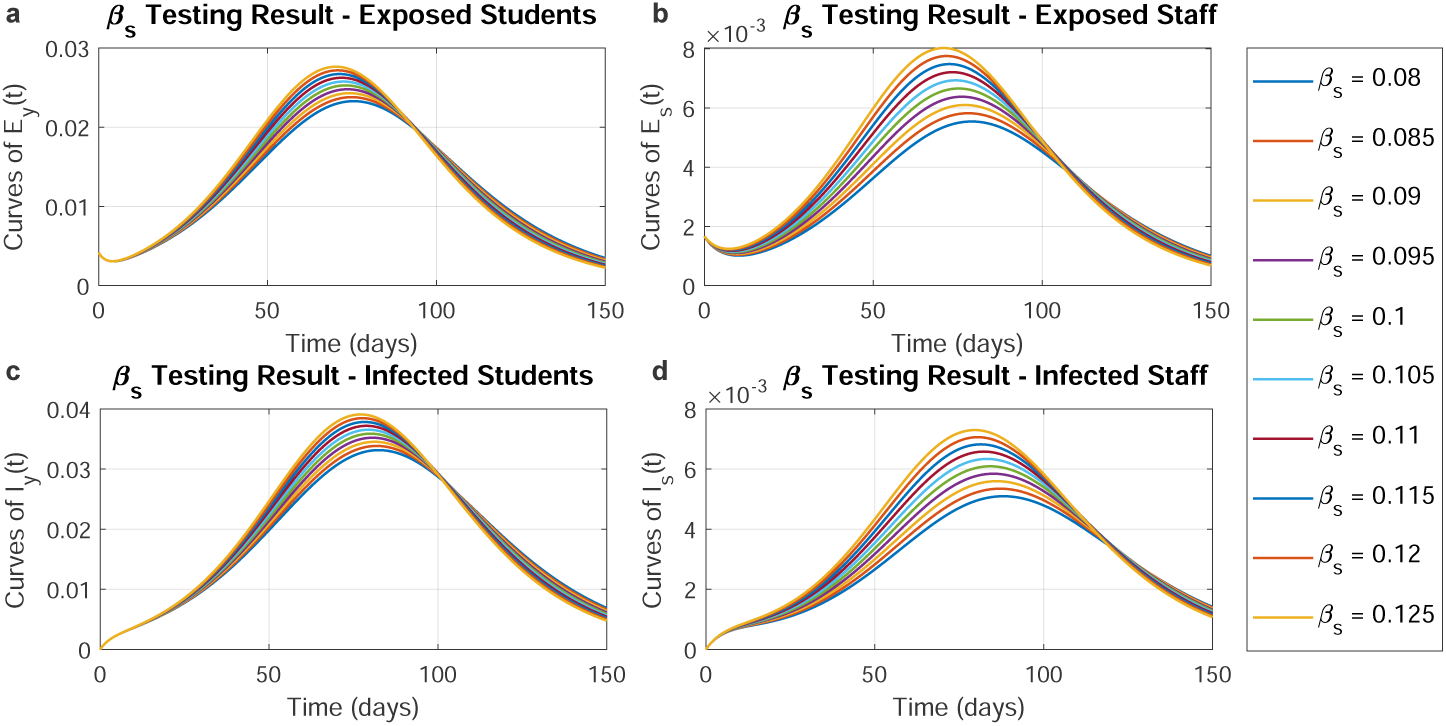
Numerical testing of student-to-student infection rate *β*_*s*_. Trajectories of cases among students and staffs when *β*_*s*_ varying from 0.08 to 0.125. With different *β*_*s*_, panels **a** and **b** show the proportion trajectories of exposed students and staffs respectively while panels **c** and **d** depict the proportion trajectories of infected students and staffs respectively.

**Fig. A3.**
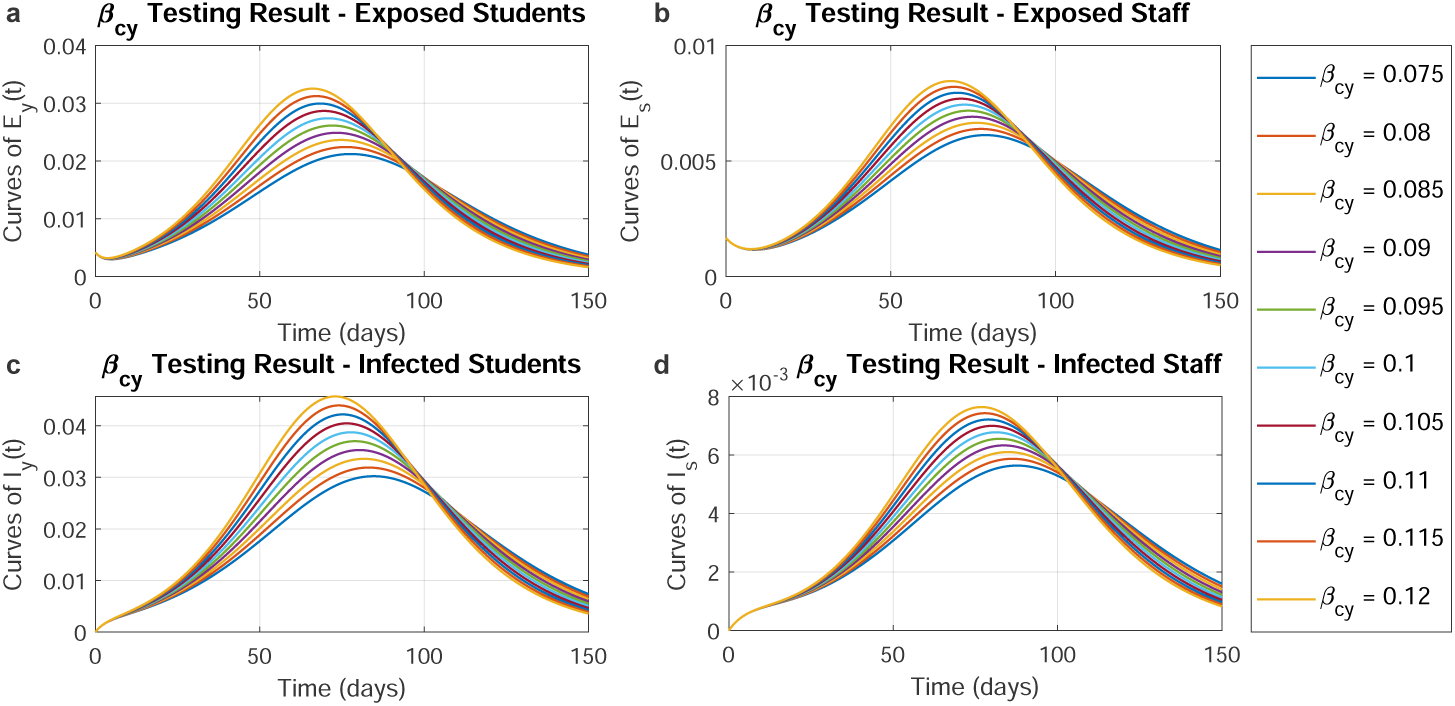
Numerical testing of student-to-student infection rate *β*_*cy*_. Trajectories of cases among students and staffs when *β*_*cy*_ varying from 0.075 to 0.12. With different *β*_*cy*_, panels **a** and **b** show the proportion trajectories of exposed students and staffs respectively while panels **c** and **d** depict the proportion trajectories of infected students and staffs respectively.

**Fig. A4.**
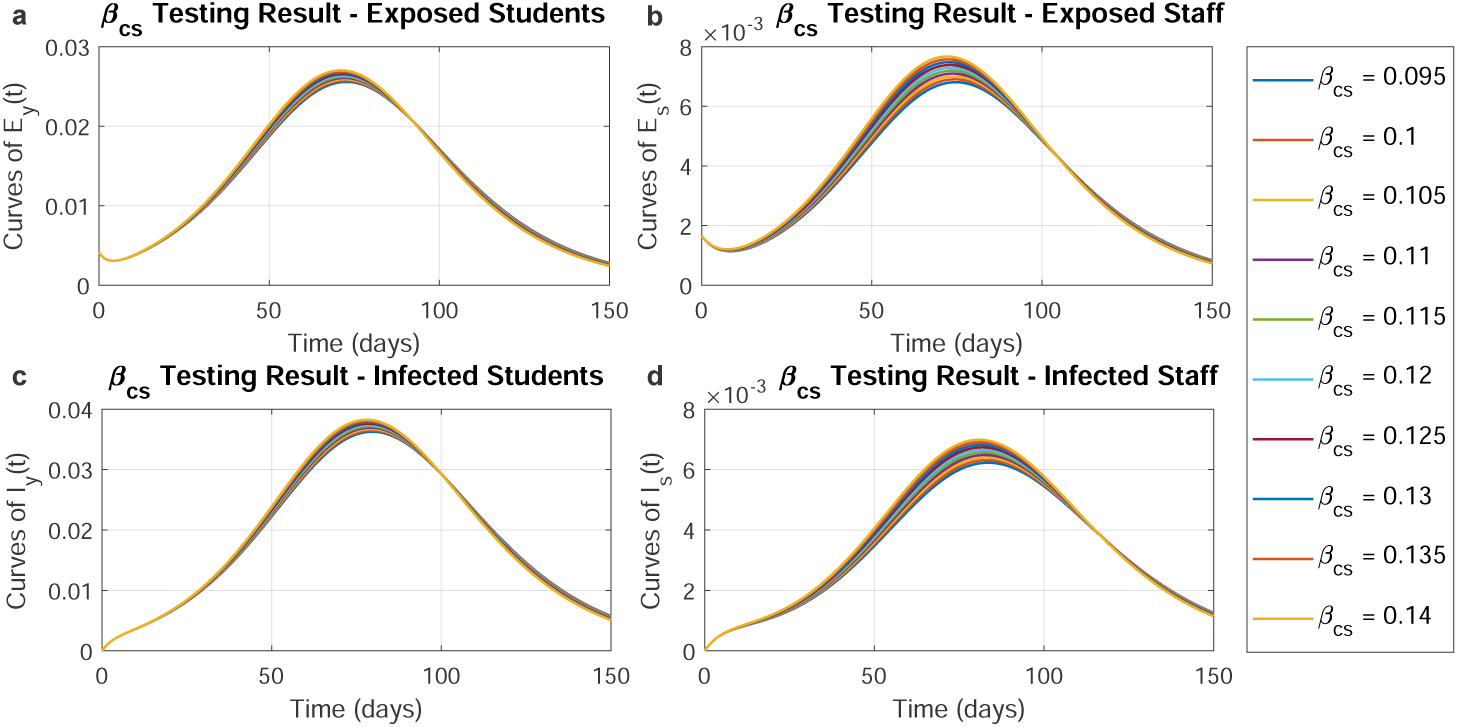
Numerical testing of student-to-student infection rate *β*_*cs*_. Trajectories of cases among students and staffs when *β*_*cs*_ varying from 0.095 to 0.14. With different *β*_*cs*_, panels **a** and **b** show the proportion trajectories of exposed students and staffs respectively while panels **c** and **d** depict the proportion trajectories of infected students and staffs respectively.

**Fig. A5.**
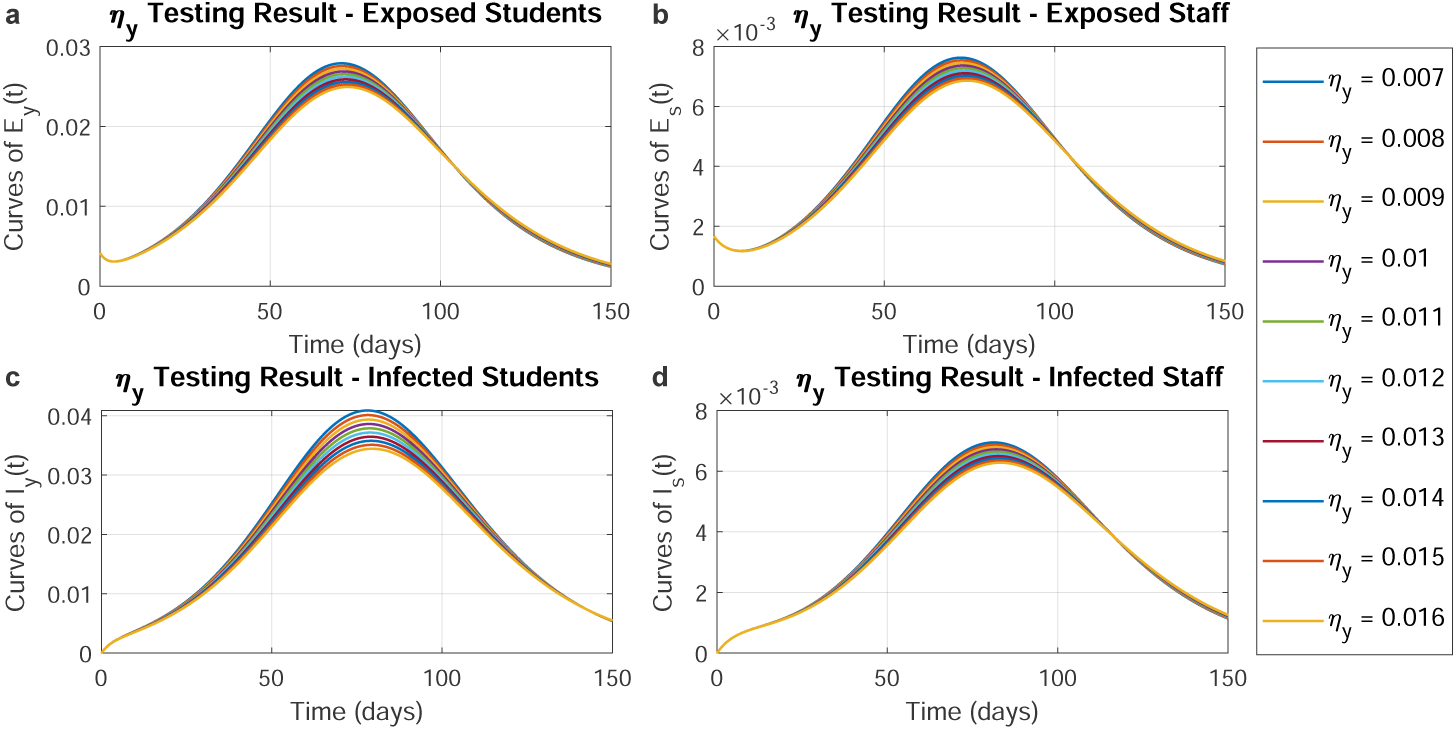
Numerical testing of student-to-student infection rate *η*_*y*_. Trajectories of cases among students and staffs when *η*_*y*_ varying from 0.007 to 0.016. With different *η*_*y*_, panels **a** and **b** show the proportion trajectories of exposed students and staffs respectively while panels **c** and **d** depict the proportion trajectories of infected students and staffs respectively.

**Fig. A6.**
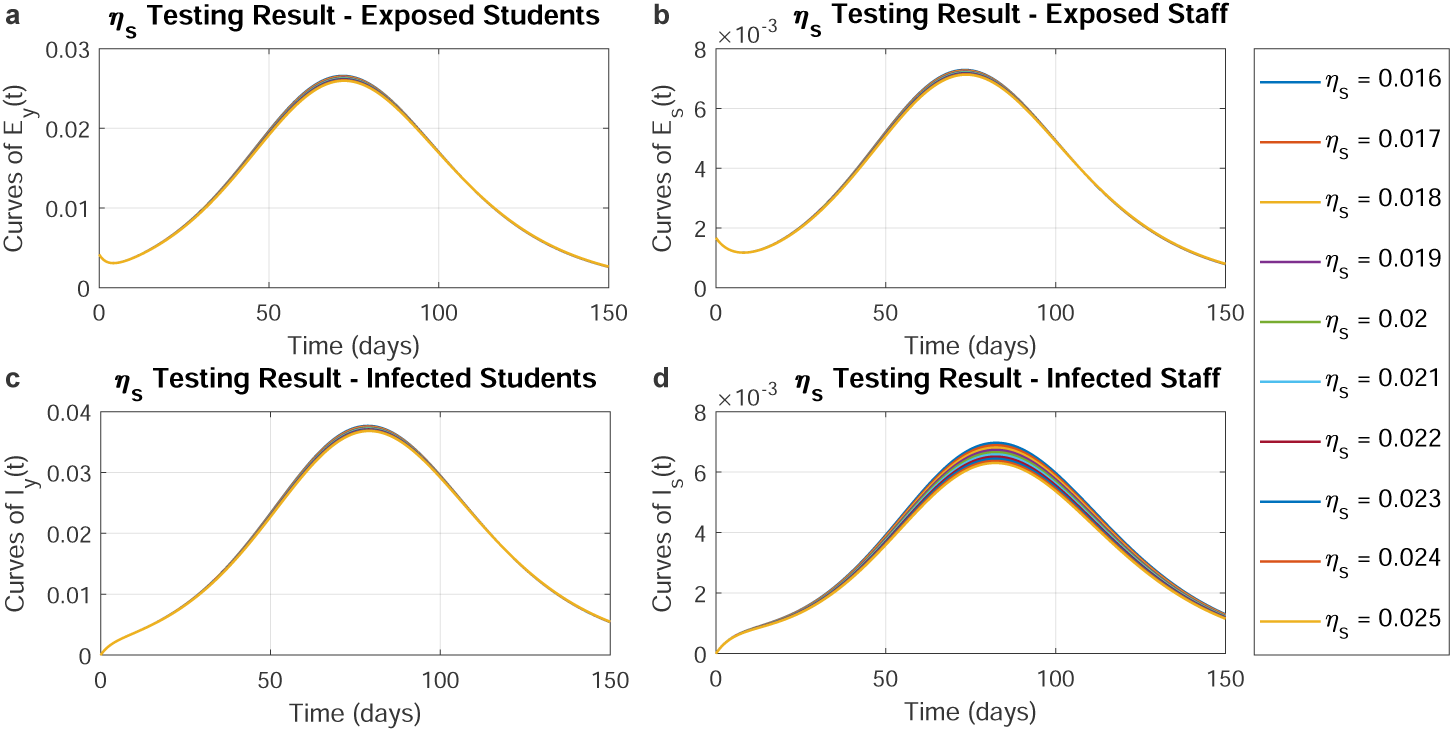
Numerical testing of student-to-student infection rate *η*_*s*_. Trajectories of cases among students and staffs when *η*_*s*_ varying from 0.016 to 0.025. With different *η*_*s*_, panels **a** and **b** show the proportion trajectories of exposed students and staffs respectively while panels **c** and **d** depict the proportion trajectories of infected students and staffs respectively.

**Fig. A7.**
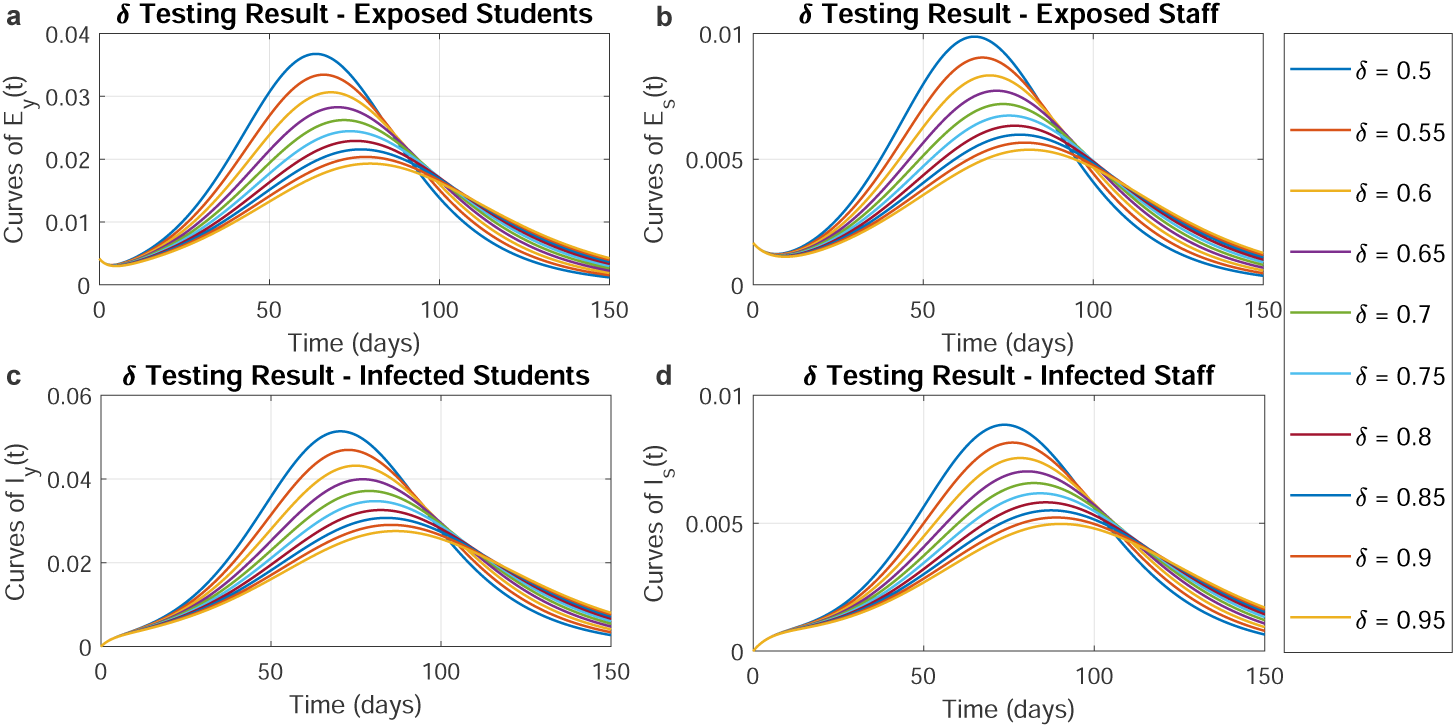
Numerical testing of student-to-student infection rate *δ*. Trajectories of cases among students and staffs when *δ* varying from 0.5 to 0.95. With different *δ*, panels **a** and **b** show the proportion trajectories of exposed students and staffs respectively while panels **c** and **d** depict the proportion trajectories of infected students and staffs respectively.

**Fig. A8.**
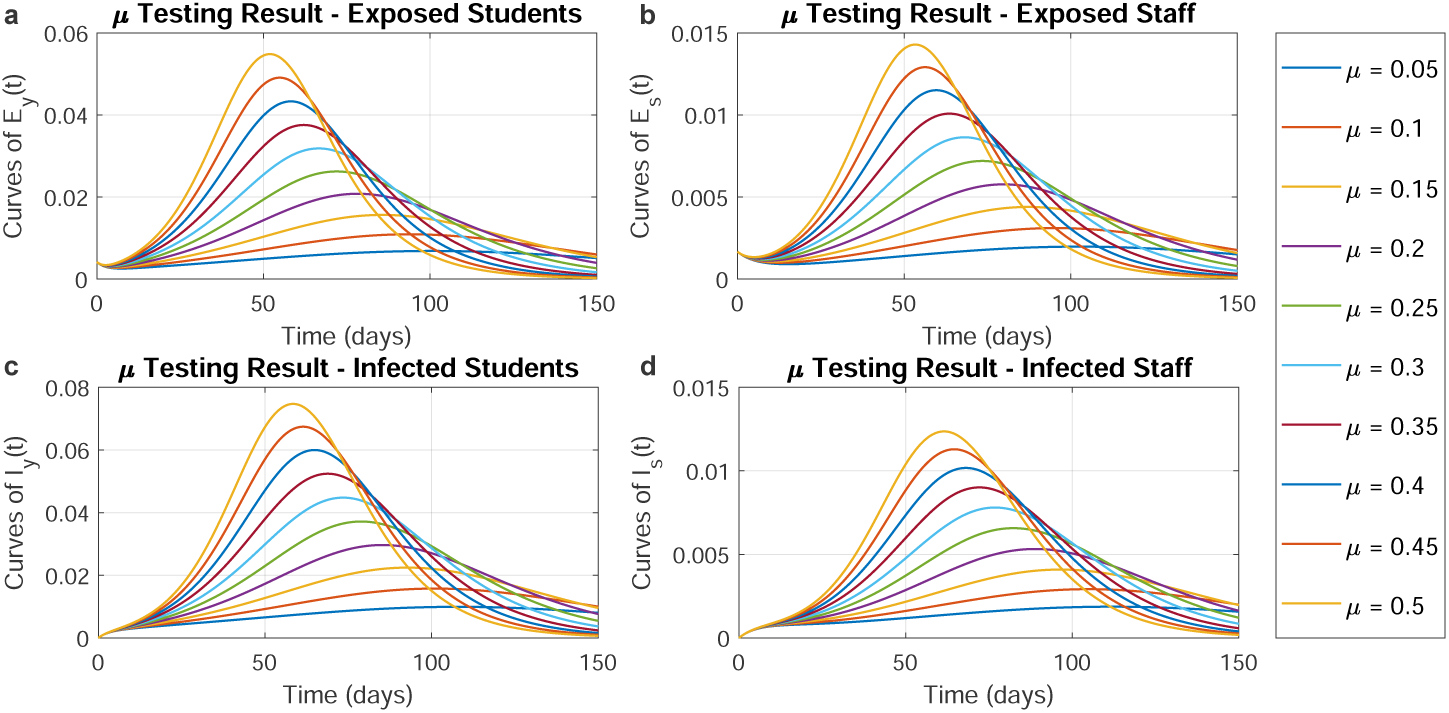
Numerical testing of student-to-student infection rate *µ*. Trajectories of cases among students and staffs when *µ* varying from 0.05 to 0.5. With different *µ*, panels **a** and **b** show the proportion trajectories of exposed students and staffs respectively while panels **c** and **d** depict the proportion trajectories of infected students and staffs respectively.

## Notes

### Competing Interest Statement

The authors have declared no competing interest.

### Funding Statement

This study did not receive any funding

## References

[1] WHO: Coronavirus disease (COVID-19) (2021). https://www.who.int/emergencies/diseases/novel-coronavirus-2019

[2] Islam, M.A., et al.: Prevalence of headache in patients with coronavirus disease 2019 (covid-19): a systematic review and meta-analysis of 14,275 patients. Frontiers in neurology 11 (2020)

[3] Agyeman, A.A., et al.: Smell and taste dysfunction in patients with covid-19: a systematic review and meta-analysis. In: Mayo Clinic Proceedings, vol. 95, pp. 1621–1631 (2020). Elsevier

[4] WHO, et al.: Modes of transmission of virus causing covid-19: implications for ipc precaution recommendations: scientific brief, 29 march 2020. Technical report, World Health Organization (2020)

[5] WHO: Getting your workplace ready for covid-19: how covid-19 spreads, 19 march 2020. Technical report, World Health Organization (2020)

[6] Qing, H., et al.: The possibility of covid-19 transmission from eye to nose. Acta ophthalmologica (2020)

[7] He, D., et al.: The relative transmissibility of asymptomatic covid-19 infections among close contacts. International Journal of Infectious Diseases 94, 145–147 (2020)

[8] Li, Y., et al.: Asymptomatic and symptomatic patients with non-severe coronavirus disease (covid-19) have similar clinical features and virological courses: a retrospective single center study. Frontiers in microbiology 11, 1570 (2020)

[9] Ritchie, H., et al.: Coronavirus pandemic (covid-19). Our World in Data (2020). https://ourworldindata.org/coronavirus

[10] Domingo, E., et al.: Basic concepts in rna virus evolution. The FASEB Journal 10(8), 859–864 (1996)

[11] Cascella, M., Rajnik, M., Aleem, A., Dulebohn, S., Di Napoli, R.: Features, evaluation, and treatment of coronavirus (covid-19). StatPearls (2021)

[12] Burki, T.: Understanding variants of sars-cov-2. The Lancet 397(10273), 462 (2021)

[13] Lopez Bernal, J., et al.: Effectiveness of covid-19 vaccines against the b.1.617.2 (delta) variant. New England Journal of Medicine 0(0), (0) https://doi.org/10.1056/NEJMoa2108891. https://doi.org/10.1056/NEJMoa2108891

[14] Lauring, A.S., Hodcroft, E.B.: Genetic variants of sars-cov-2—what do they meanã Jama 325(6), 529–531 (2021)

[15] Kimura, I., et al.: Sars-cov-2 lambda variant exhibits higher infectivity and immune resistance. Biorxiv (2021)

[16] Weisblum, Y., et al.: Escape from neutralizing antibodies by sars-cov-2 spike protein variants. Elife 9, 61312 (2020)

[17] Greaney, A.J., et al.: Complete mapping of mutations to the sars-cov-2 spike receptor-binding domain that escape antibody recognition. Cell host & microbe 29(1), 44–57 (2021)

[18] Israel, A., et al.: Large-scale study of antibody titer decay following bnt162b2 mrna vaccine or sars-cov-2 infection. medRxiv (2021)

[19] Ibarrondo, F.J., et al.: Primary, recall, and decay kinetics of sars-cov-2 vaccine antibody responses. ACS nano (2021)

[20] Fontanet, A., et al.: Sars-cov-2 variants and ending the covid-19 pandemic. The Lancet 397(10278), 952–954 (2021)

[21] Cooper, I., Mondal, A., Antonopoulos, C.G.: A sir model assumption for the spread of covid-19 in different communities. Chaos, Solitons and Fractals 139, 110057 (2020). https://doi.org/10.1016/j.chaos.2020. 110057

[22] Leontitsis, A., et al.: Seahir: A specialized compartmental model for covid-19. International Journal of Environmental Research and Public Health 18, 2667 (2021). https://doi.org/10.3390/ijerph18052667

[23] Giordano, G., et al.: Modelling the covid-19 epidemic and implementation of population-wide interventions in italy. Nature medicine 26(6), 855–860 (2020)

[24] Aguiar, M., Ortuondo, E.M., Van-Dierdonck, J.B., Mar, J., Stollenwerk, N.: Modelling covid 19 in the basque country from introduction to control measure response. Scientific reports 10(1), 1–16 (2020)

[25] Abrams, S., et al.: Modelling the early phase of the belgian covid-19 epidemic using a stochastic compartmental model and studying its implied future trajectories. Epidemics 35, 100449 (2021)

[26] Chatterjee, K., Chatterjee, K., Kumar, A., Shankar, S.: Healthcare impact of covid-19 epidemic in india: A stochastic mathematical model. Medical Journal Armed Forces India 76(2), 147–155 (2020)

[27] Leach, A., Clarke, S., Kirk, A.: Covid UK: coronavirus cases, deaths and vaccinations today (2021). http://www.theguardian.com/world/2021/aug/20/covid-uk-coronavirus-cases-deaths-and-vaccinations-today

[28] Iacobucci, G.: Covid-19: Infections fell by 65% after first dose of astrazeneca or pfizer vaccine, data show. bmj 373, 1068 (2021)

[29] Menni, C., et al.: Vaccine side-effects and sars-cov-2 infection after vaccination in users of the covid symptom study app in the uk: a prospective observational study. The Lancet Infectious Diseases (2021)

[30] COVID-19 Pandemic Planning Scenarios (2021). https://www.cdc.gov/coronavirus/2019-ncov/hcp/planning-scenarios.html

[31] Interim Clinical Guidance for Management of Patients with Confirmed Coronavirus Disease (COVID-19) (2021). https://www.cdc.gov/coronavirus/2019-ncov/hcp/clinical-guidance-management-patients.html

[32] Lopman, B., et al.: A model of covid-19 transmission and control on university campuses. medRxiv (2020). https://doi.org/10.1101/2020.06.23.20138677

[33] Voinsky, I., Baristaite, G., Gurwitz, D.: Effects of age and sex on recovery from covid-19: Analysis of 5769 israeli patients. Journal of Infection 81(2), 102–103 (2020)

[34] George, N., Tyagi, N.K., Prasad, J.B.: Covid-19 pandemic and its average recovery time in indian states. Clinical Epidemiology and Global Health 11, 100740 (2021). https://doi.org/10.1016/j.cegh.2021.100740

[35] Van Doremalen, N., et al.: Aerosol and surface stability of sars-cov-2 as compared with sars-cov-1. The New England journal of medicine 382(16), 1564–1567 (2020). https://doi.org/10.1056/nejmc2004973

[36] Anderson, E.L., Turnham, P., Griffin, J.R., Clarke, C.C.: Consideration of the aerosol transmission for covid-19 and public health. Risk Analysis 40(5), 902–907 (2020)

[37] Fiorillo, L., et al.: Covid-19 surface persistence: a recent data summary and its importance for medical and dental settings. International journal of environmental research and public health 17(9), 3132 (2020)

[38] Moghadas, S.M., et al.: The impact of vaccination on coronavirus disease 2019 (covid-19) outbreaks in the united states. Clinical Infectious Diseases (2021)

[39] Miller, E.: 1 - global burden of disease: Part c. potential and existing impact of vaccines on disease epidemiology. In: Bloom, B.R., Lambert, P.-H. (eds.) The Vaccine Book, pp. 37–50. Academic Press, San Diego (2003). https://doi.org/10.1016/B978012107258-2/50005-6. https://www.sciencedirect.com/science/article/pii/B9780121072582500056

[40] Driessche, P., Watmough, J.: Further Notes on the Basic Reproduction Number, vol. 1945, pp. 159–178 (2008). https://doi.org/10.1007/978-3-540-78911-66

[41] Van Den Driessche, P., Watmough, J.: Reproduction numbers and sub-threshold endemic equilibria for compartmental models of disease transmission. Mathematical Biosciences 180(1), 29–48 (2002). https://doi.org/10.1016/S0025-5564(02)00108-6

[42] Diekmann, O., Heesterbeek, J., Roberts, M.G.: The construction of next-generation matrices for compartmental epidemic models. Journal of the Royal Society Interface 7(47), 873–885 (2010)

[43] Karaivanov, A., Lu, S.E., Shigeoka, H., Chen, C., Pamplona, S.: Face masks, public policies and slowing the spread of covid-19: Evidence from canada. medRxiv (2020). https://doi.org/10.1101/2020.09.24.20201178

[44] Jarvis, C.I., et al.: Quantifying the impact of physical distance measures on the transmission of covid-19 in the uk. BMC medicine 18(1), 1–10 (2020)

[45] Lindsley, W.G., Blachere, F.M., Law, B.F., Beezhold, D.H., Noti, J.D.: Efficacy of face masks, neck gaiters and face shields for reducing the expulsion of simulated cough-generated aerosols. Aerosol Science and Technology 55(4), 449–457 (2021). https://doi.org/10.1080/02786826.2020.1862409

[46] Ueki, H., et al.: Effectiveness of face masks in preventing airborne transmission of sars-cov-2. mSphere 5(5), 00637–20 (2020). https://doi.org/10.1128/mSphere.00637-20

